# Partnership-based sexual behaviours among adults in Singapore

**DOI:** 10.1101/2025.09.16.25335841

**Authors:** A Janhavi, Gregory Gan, Akira Endo, Rayner Kay Jin Tan, Natasha Howard, Kiesha Prem, Michelle Ho, Shihui Jin, Borame L Dickens

## Abstract

**Introduction:** Sexual behaviour data are essential for monitoring behavioural trends and designing effective sexual health interventions. However, in Singapore and Southeast Asia more broadly, most research has focused on key populations, leaving a decades-long research gap in the general adult population. To address this, characterised sexual behaviour patterns and examined associations with socio-demographic characteristics, engagement with sex workers, and STI positivity.

**Methods:** We conducted a nationwide cross-sectional online survey (September 2023 to July 2024). Of 2,876 respondents, 2,297 were eligible (aged 21 years and above with complete key demographic data). Participants reported socio-demographic characteristics and sexual behaviours within the past year, 6 months, and 3 months. Outcomes included fitted distributions of partner numbers across partnered activities (mutual masturbation, oral, vaginal, anal sex) and associations with socio-demographic factors, sex worker engagement, and self-reported STI positivity.

**Results:** Of participants, 51.5% were male, 84.4% were Singapore citizens, 69.6% were aged between 21-60. Individuals reporting sexual partners of the same sex had a higher probability of having 5 or more same-sex partners respectively in the past year (WSW: 28.4% mutual masturbation, 27.8% oral sex; MSM: 36.2% mutual masturbation, 34.5% oral sex). Engagement with sex workers was reported by 4.7% (95% CI, 3.7-6.2%) of females and 18.1% (16.1-20.4%) of males, with higher odds among individuals with at least 5 partners in the past year (e.g., vaginal sex AOR = 7.40, 95% CI, 4.03-13.58). Self-reported STI positivity was 13.1%, associated with having at least 5 partners in the past year (e.g., vaginal sex AOR = 5.03, 95% CI, 1.44-17.61) and ever engaging with sex workers (e.g., vaginal sex AOR = 3.16, 95% CI, 1.67-5.99), but not with barrier protection frequency.

**Conclusions:** Patterns varied by sex, sexual orientation, and risk behaviours, underscoring the need for context-specific behavioural data to inform STI modelling and public health interventions.

## Introduction

Understanding sexual health and behaviour is crucial for promoting overall public health, as it has profound implications on both physical^1^ and mental health^2^, and shapes health outcomes for sexually transmitted infections (STIs)^3^. However, cultural norms, societal values, and religious beliefs in many parts of Asia often inhibit open dialogue about sexual matters^4,5^. Against this backdrop, research efforts have often concentrated on specific key populations, while the general adult population remains comparatively understudied, particularly in terms of the depth of detailed sexual behaviour data^6^.

This pattern is also evident in Singapore, where research on adult sexual health and behaviour has predominantly focused on key populations, such as men who have sex with men (MSM)^7,8^, sex workers^9–11^, and heterosexual male clients of sex workers^12,13^. These studies have largely focused on STIs and participation in risky sexual behaviours. High-risk adolescents have also been studied, particularly in the context of sexual education and STI prevention^14,15^. In contrast, sexual behaviour within the general population remains underexplored. The last published population-level sexual behaviour survey was conducted in the 1990s^16^. Although more recent studies have included general population samples, they have largely focused on changes in sexual behaviour during the COVID-19 pandemic or psychosocial factors such as stress and fatigue^17–19^. While these studies offer valuable insight, they are limited in scope, lacking detailed information on the frequency, variety, and demographic differences of specific sexual behaviours.

The lack of comprehensive, contemporary data limits our understanding of the frequency and variety of sexual behaviours among adults in Singapore. As cultural norms around sexual behaviour shift, attitudes toward sex likely vary across age groups, with younger populations potentially being more sexually active^20,21^. Yet, this dynamic remains poorly characterized in Singapore. Apart from age, diversity in sexual behaviour across other demographic groups, such as ethnicity and sexual orientation, also remain under-characterized. Moreover, detailed information on specific sexual behaviours – such as how often different types of partnered sexual activity (e.g., mutual masturbation, oral, vaginal, anal sex) occur, how they differ across time frames (e.g., 1 year, 6 months, 3 months), if they occur with different partners, and whether barrier protection (e.g., condoms or dental dams) is used across different sexual acts – is lacking.

To fill this knowledge gap, we conducted an online survey to characterize sexual behaviours in Singapore’s general adult population. Specifically, we aimed to: (i) describe sexual behaviour patterns by estimating the fitted distributions of the number of sexual partners across different partnered sexual activities, stratified by sex, sexual partner preference, and recall period (1 year, 6 months, 3 months); investigate associations between sexual behaviours and socio-demographic characteristics, and (ii) engagement with sex workers, or (iii) self-reported STI positivity. These findings would inform targeted sexual health interventions and public health messaging in Singapore and the broader Southeast Asian region.

## Methods

### Survey design and data collection

Between September 2023 and July 2024, an online survey on sexual behaviour was conducted in English in Singapore. Responses were collected through multiple recruitment strategies to reach diverse demographic groups. To engage younger participants, the survey was shared on social media platforms that are widely used by younger demographics (Facebook and Telegram). To increase the representation of individuals with diverse sexual orientations, the survey was promoted in collaboration with Pink Dot SG, a non-profit movement supporting the LGBTQ community, via their Instagram account and at the offline Pink Dot 16 event. To reach older residents, responses were also gathered through the survey company IPSOS. All participants provided informed consent and completed the survey voluntarily. Both surveys ran simultaneously.

In the survey, respondents first answered multiple-choice questions on their socio-demographics, such as age, sex assigned at birth (male/female), ethnicity, religion, resident status within Singapore, and sexual identity. They were also asked to disclose the sex assigned at birth of their past and current sexual partners. Respondents were excluded from completing the survey if they were under 21 years old (as required by ethics), refused to disclose their age, sex assigned at birth of themselves and/or their sexual partners (if any), or resided outside of Singapore in the past six months. As a part of quality control, a reCAPTCHA verification was placed at the start of the survey to prevent automated or non-human entries, where only those who passed verification could proceed to the questionnaire. Among the remaining eligible respondents, those who reported having at least one sexual partner in their lifetime were then prompted with follow-up questions on their sexual behaviours with either or both sexes. These questions covered the number of sexual partners, frequency of sexual activity, and barrier protection usage for each of the four partnered sexual activities, including mutual masturbation, oral sex, vaginal sex, and anal sex. Participants were also asked about sex toy usage and engagement with sex workers (i.e., those who receive money or goods in exchange for sexual services). The recall periods for these questions generally included past year, past six months, and past three months. In addition, the survey also assessed STI-related factors, including STI positivity, testing frequency, and symptoms and treatments received for those claiming to be STI-positive. Please refer to Supplementary Information for the full survey and further details on data collection.

### Statistical analysis

We summarized the socio-demographic characteristics and sexuality for all eligible participants using counts and percentages. Individuals who reported having sexual partners but indicated no engagement in partnered sexual activities were recoded as having no sexual partners. For the frequency of sexual behaviours, responses such as “Not sure”, “Too many to count”, or “Lost track” were treated invalid and missing. When a range of values were provided, the mean of the upper and lower bounds was used. If only a single value was given, it was recorded as is.

To characterize sexual contact patterns in Singapore, we focused on the four sexual activities (mutual masturbation, oral sex, vaginal sex, and anal sex), estimating the proportion of individuals aged 21–60 who engaged in each activity. Meanwhile, among those who reported having at least one sexual partner, we approximated the number of sexual partners and the frequency of sexual activities utilizing a Lomax distribution to account for the heavy-tailed nature of the observations. Specifically, let *N* > 0 denote the number of partners and *n* the realization. The probability of having more than *n* partners (i.e., at least (*n* + 1) partners) is expressed as

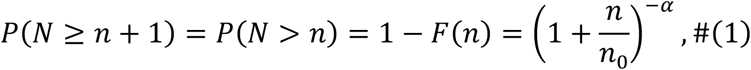

where *F*(⋅) is the cumulative density function of *N*, and *n*_0_ and α are positive parameters to be estimated. In addition, the distribution of the number of times a survey participant engaged in sexual activities was also modelled in the format of (1). We defined four partnership types based on the sex assigned at birth of both the survey participants and their partners, including female respondents with female partners, female respondents with male partners, male respondents with female partners, and male respondents with male partners. Individuals with partners of both sexes were classified into two categories simultaneously. We fitted the model to all combinations of the four partnership types, three recall periods, and four sexual activity types included in the survey. We also estimated the prevalence of sex worker engagement and the distribution of the number of sex workers one engaged with among individuals who reported engaging with them. The detailed estimation approach can be found in the Supplementary Information.

We further constructed logistic regression models to investigate the relationship between socio-demographics, sexual behaviour patterns, and STI positivity among individuals aged 21–60. As with the previous analysis, the respondents were also classified based on the sex assigned at their birth. For those who reported having sexual partners in the past year, their partnership-based sexual orientation was determined by the disclosed sex assigned at birth of their partner(s), while for those without sexual partners, the variable was determined based on their sex and the self-reported sexual orientation. In total, we included seven distinct partnership-based sexual orientations: men who have sex with men only (MSMO), men who have sex with women only (MSWO), men who have sex with both sexes (MSMW), women who have sex with men only (WSMO), women who have sex with women only (WSWO), women who have sex with both sexes (WSMW), and others. We considered three binary variables as the outcome variable in the regression, including i) sexual partnership status (i.e, whether to engage in a specific type of sexual activity with one or more sexual partners) in the past year, ii) engagement with sex workers, and iii) reporting any STI positivity. The predictors encompassed a subset of age, ethnicity, partnership-based sexual orientation, number of sexual partners in the last year, engagement with sex workers, STI positivity, and barrier protection usage frequency. See Supplementary Information for detailed descriptions of the models (Supplementary Table 1). Adjusted odds ratios were estimated using the survey package in R^22^.

All data were assumed to be missing at random, with no imputation applied. Sex was defined based on the sex assigned at birth unless otherwise stated. For the analyses of sexual contact patterns and their association with socio-demographics and sexual behaviour patterns, we applied sample weights based on the age and sex composition of Singaporean residents^23^ to improve the representativeness of our estimates for the general population in Singapore. All the analyses were performed using the R software^24^.

### Ethics

Ethics approval was obtained from the NUS Institutional Review Board (reference NUS-IRB-2023-377).

## Results

### Participant characteristics

A total of 2,297 participants were included in the analysis after exclusion (n=579; details in Methods), with their socio-demographics and sexuality summarized in Table 1. Approximately half of the participants were males (51.5%). The majority were Singapore citizens (84.4%), aged 21–40 (69.6%), of Chinese ethnicity (59.6%), and heterosexual (77.3%). A higher proportion of female respondents reported having no sexual partners compared to males (Table 1). While the response rates for the socio-demographic questions were generally high, those for certain sensitive questions (e.g., STI positivity) fell below 80% in specific age groups (Supplementary Table 2).

**Table 1.** Socio-demographics and sexuality of all eligible participants, stratified by sex assigned at birth. signed at birth.

|  | **Males** |  | **Females** |  |
| --- | --- | --- | --- | --- |
|  | 51.5% (N = 1183) |  | 48.5% (N = 1114) |  |
|  | % | N | % | N |
| **Age** |  |  |  |  |
| 21–30 | 16.2 | 373 | 20.5 | 472 |
| 31–40 | 15.6 | 359 | 17.3 | 397 |
| 41–50 | 9.9 | 227 | 6.3 | 144 |
| 51–60 | 6.3 | 145 | 3.4 | 78 |
| 61+ | 3.4 | 79 | 1.0 | 23 |
| 21–30 | 16.2 | 373 | 20.5 | 472 |
| **Gender** |  |  |  |  |
| Man | 50.7 | 1165 | 0.9 | 21 |
| Woman | 0.4 | 9 | 46.9 | 1078 |
| Others | 0.4 | 9 | 0.7 | 15 |
| **Ethnicity** |  |  |  |  |
| Chinese | 35.5 | 815 | 24.1 | 553 |
| Malay | 6.9 | 159 | 11.5 | 265 |
| Indian | 7.2 | 165 | 12.4 | 284 |
| Others | 1.6 | 36 | 0.5 | 11 |
| Unknown | 0.3 | 8 | 0.0 | 1 |
| **Residential Status** |  |  |  |  |
| Singapore citizen | 44.0 | 1010 | 40.4 | 928 |
| Permanent resident | 4.9 | 113 | 6.5 | 149 |
| Work pass holder | 2.1 | 48 | 1.0 | 24 |
| Others | 0.3 | 7 | 0.4 | 11 |
| Unknown | 0.2 | 5 | 0.1 | 2 |
| **Religion** |  |  |  |  |
| Non-religious<sup>a</sup> | 8.7 | 199 | 6.7 | 154 |
| Buddhism | 12.3 | 282 | 9.5 | 218 |
| Christianity | 11.1 | 256 | 9.6 | 221 |
| Hinduism | 4.4 | 102 | 7.6 | 175 |
| Islam | 7.3 | 168 | 11.1 | 255 |
| Taoism | 3.0 | 68 | 2.2 | 51 |
| Others | 0.3 | 6 | 0.1 | 2 |
| Unknown | 4.4 | 102 | 1.7 | 38 |
| **Monthly salary** |  |  |  |  |
| No income | 7.1 | 162 | 6.8 | 156 |
| SGD0–2000 | 4.4 | 101 | 6.6 | 151 |
| SGD2000–4999 | 14.8 | 341 | 17.5 | 401 |
| SGD5000–7999 | 12.1 | 277 | 9.8 | 225 |
| SGD8000–9999 | 4.7 | 108 | 4.2 | 96 |
| SGD10000+ | 6.8 | 157 | 3.1 | 71 |
| Unknown | 1.6 | 37 | 0.6 | 14 |
| **Sexual identity** |  |  |  |  |
| Heterosexual | 35.7 | 820 | 41.6 | 956 |
| Homosexual | 9.7 | 223 | 0.8 | 19 |
| Bisexual | 4.0 | 92 | 3.3 | 76 |
| Questioning | 0.7 | 17 | 1.0 | 22 |
| Others | 0.3 | 8 | 0.7 | 15 |
| Queer | 0.2 | 5 | 0.7 | 17 |
| Unknown | 0.8 | 18 | 0.4 | 9 |
| **Partner's sex** |  |  |  |  |
| Female | 29.9 | 687 | 2.9 | 66 |
| Male | 10.4 | 238 | 30.4 | 699 |
| Both | 2.8 | 65 | 1.5 | 34 |
| No sexual partners<sup>b</sup> | 8.4 | 193 | 13.7 | 315 |

<sup>a</sup>Including individuals who were non-religious, atheist, agnostic, or freethinkers.
<sup>b</sup>Including individuals who had not participated in any of the four partnered sexual activities.

### Self-reported sexual activity

Compared to females, males were more likely to report having engaged in mutual masturbation and oral sex. While vaginal sex was relatively uncommon among MSM, with 13.0% (95% confidence interval [CI]: 0–39.0%) reporting least one such partner in the past year, this group had the highest likelihood to engage in anal sex (48.6%, 95% CI: 43.7%– 55.3%) across different subpopulations. In contrast, women who have sex with women (WSW) were the least likely to engage in all the sexual activities except for vaginal sex, with fewer than 25% reporting one or more sexual partners for the respective activities (Figure 1, Supplementary Figure 8–10).

**Figure 1:**
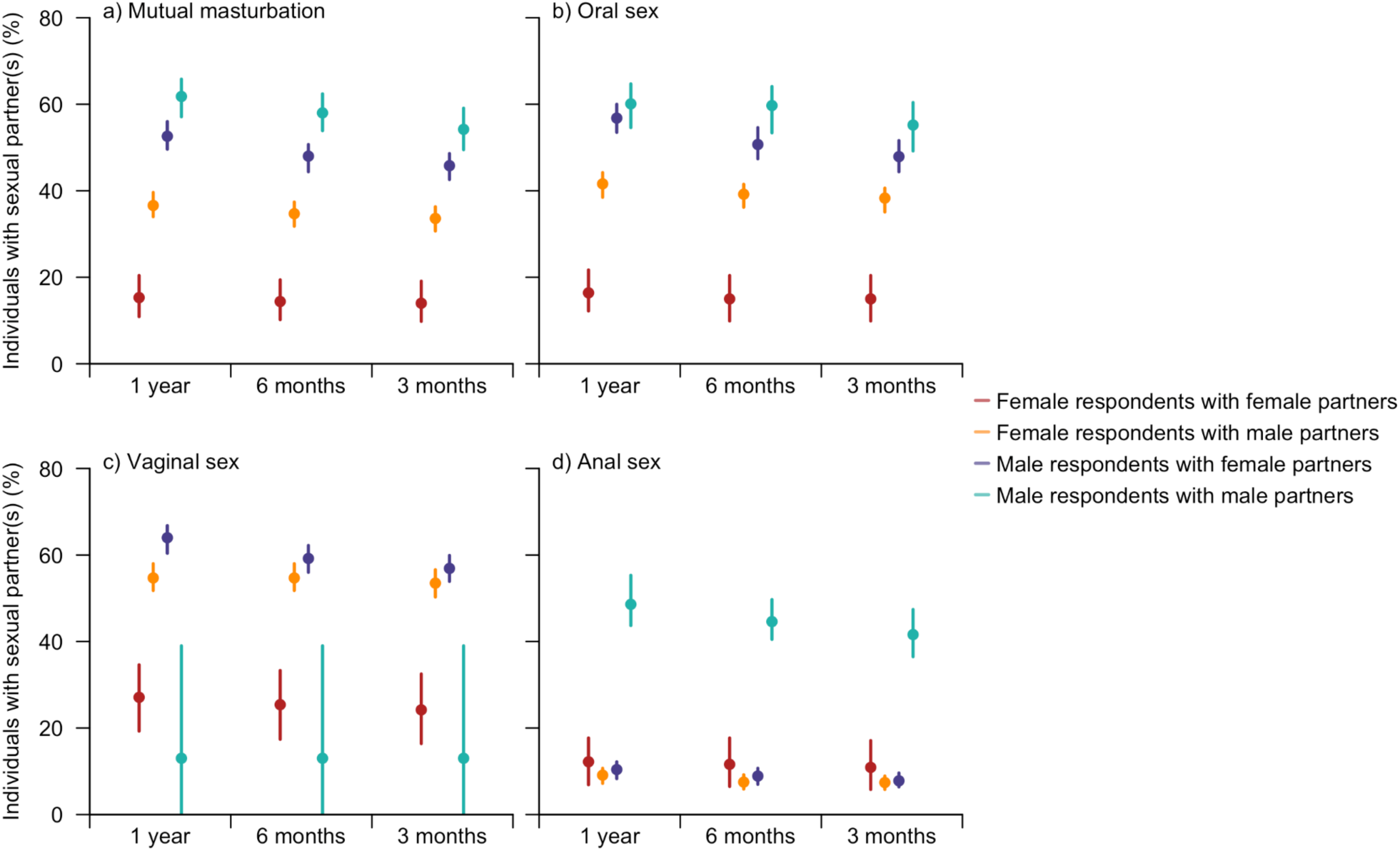
Estimated proportion of individuals with at least one sexual partner, stratified by the sex assigned at birth of survey respondents and their partners (for the specified sexual activity), type of sexual activity, and recall period. Dots represent estimated mean values and lines indicate the corresponding 95% CIs. Female or male respondents with partners of both sexes were classified into two categories simultaneously.

Nevertheless, among individuals with at least one sexual partner, WSW were the most likely to have high numbers of partners compared to other subpopulations. For instance, the probability of WSW having five female partners or more was estimated at 28.4% for mutual masturbation, 27.8% for oral sex, 34.6% for vaginal sex, and 71.2% for anal sex. The distribution for the number of male partners among MSM was also heavy-tailed, with the probability of having five partners or more being 36.2% for mutual masturbation and 34.5% for oral sex (Figure 2). Compared to the number of partners, differences in the frequency of sexual activities across various partner sex combinations were less pronounced, especially for mutual masturbation and oral sex (Supplementary Figure 1, Supplementary Figure 11–13).

**Figure 2:**
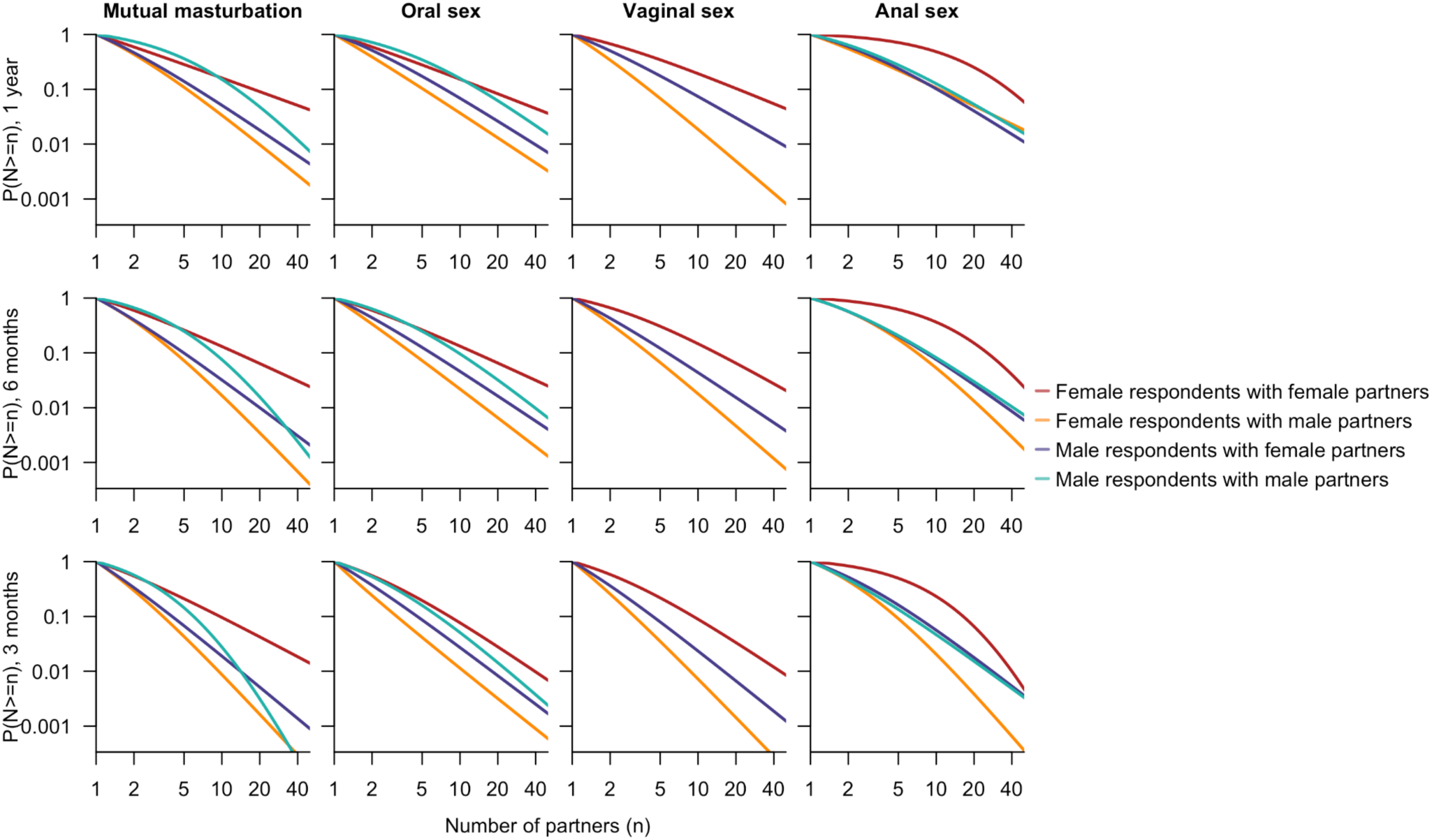
Fitted distribution of the number of partners (N) among individuals with sexual partners, stratified by the sex assigned at birth of survey respondents and their partners, type of sexual activity (columns), and recall period (rows). Female or male respondents with partners of both sexes were classified into two categories simultaneously. The distribution for the number of male partners among male respondents who engaged in vaginal sex with males was not fitted or presented due to the small sample size. Both the x- and y-axes are displayed on a logarithmic scale.

Age emerged as another factor affecting engagement in each of the four types of sexual activities (Supplementary Figure 2). Compared to 21–30, individuals aged 31–40 were more likely to have vaginal sex (adjusted odds ratio [AOR]: 1.57, 95% CI: 1.22–2.02). By contrast, people aged over 40 years were less sexually active in the other three types of sexual activities. Among those aged 41–50 years, the adjusted odds ratio of having at least one sexual partner in the past year was estimated at 0.55 (95% CI: 0.41–0.73) for mutual masturbation, 0.75 (95% CI: 0.56–1.00) for oral sex, and 0.57 (95% CI: 0.37–0.87) for anal sex. Even lower levels were estimated for the 51–60 group, with adjusted odds ratios of 0.31 (95% CI: 0.22–0.44), 0.42 (95% CI: 0.30–0.59), and 0.31 (95% CI: 0.17–0.59), respectively, for the aforementioned three sexual activities. These two groups also had a substantially lower likelihood to have at least five partners across the three types of sexual activity in the past year (Supplementary Figure 3).

### Engagement with sex workers

Partnership-based sexual orientation was inferred to be highly correlated with engagement with sex workers, where women who have sex with men were consistently the least likely to engage with sex workers across all four partnered sexual activities (Figure 3). Overall, females were significantly less likely than males to engage in sex workers, with an estimated 4.7% (95% CI: 3.7%–6.2%) reporting such engagement in the past year. This proportion rose to 18.1% (95% CI: 16.1%–20.4%) among males (Supplementary Figure 4). These estimates slightly decreased with a shorter recall period. Females were also less likely to engage with high numbers of sex workers compared to males (Supplementary Figure 5). Apart from sex, a strong association was also observed between engagement with sex workers and sexual activity level, quantified through the number of sexual partners in the past year, across all sexual activity types. Within each group reporting mutual masturbation, oral sex, vaginal sex, and anal sex within the past year, individuals who had at least five sexual partners had significantly higher odds of engaging with sex workers compared to those with no sexual partners. The adjusted odds ratios were 4.67 (95% CI: 2.83–7.73), 6.96 (95% CI: 4.16–11.64), 7.40 (95% CI: 4.03–13.58), and 11.37 (95% CI: 5.50–23.53), respectively (Figure 3). Similar trends were observed for these factors in their impacts on frequent engagement with sex workers (Supplementary Figure 6).

**Figure 3:**
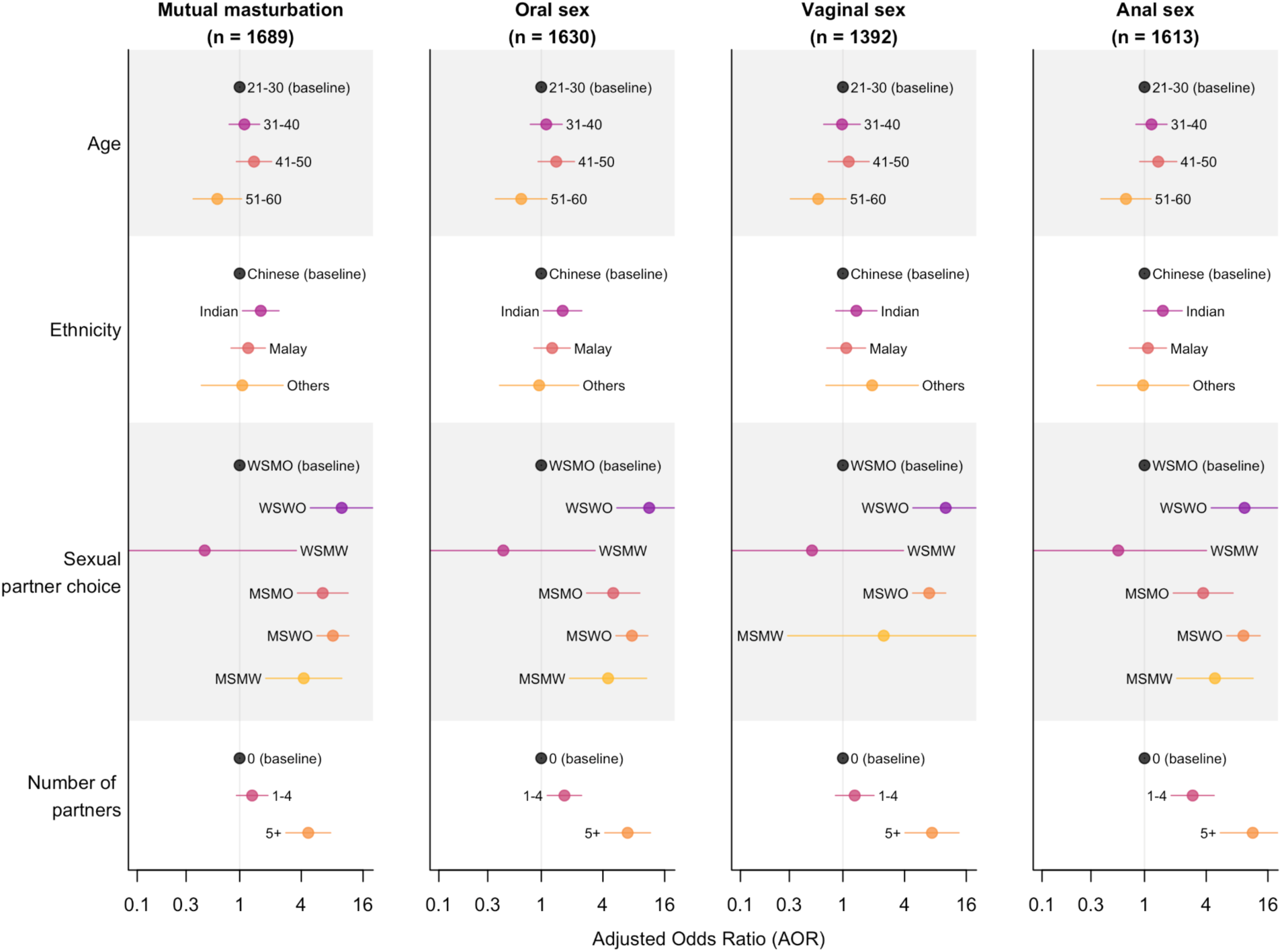
Adjusted odds ratio for socio-demographics, partnership-based sexual orientation defined by sex assigned at birth, and number of sexual partners in the past year, stratified by sexual activity type (columns). These include point estimates (dots) and corresponding 95% CIs (lines), quantifying the factors’ impacts on engagement with sex workers. Extreme estimates due to insufficient samples (e.g., vaginal sex among MSMO) are not shown. Please also note the x-axes are presented on a logarithmic scale.

### STI positivity

In total, 218 (13.1%) respondents aged 21–60 reported being diagnosed or tested positive for at least one STI, with nearly half (105) aged 21–30, and a negative correlation between positivity and age (Supplementary Table 3). However, after adjusting for testing frequency and partnership-based sexual orientation, the association between STI positivity and age was not statistically significant.

While men who have sex with women tended to have lower odds of reporting STI-positivity than other groups when other conditions were adjusted for, higher odds were associated with having at least five sexual partners, engaging with sex workers, and undergoing STI testing once every three or six months. Particularly, having at least five sexual partners over the past year was correlated with an increase in the odds of STI positivity 2.60 (95% CI: 1.22–5.54) times for mutual masturbation, 4.04 (95% CI: 1.55–

10.54) times for oral sex, 5.03 (95% CI: 1.44–17.61) times for vaginal sex, and 4.52 (95% CI: 2.22–9.20) times for anal sex among individuals ever tested for STIs and engaged in respective sexual activities. Similarly, having ever engaged with sex workers led to 3.99 (95% CI: 2.14–7.43), 3.61 (95% CI: 1.94–6.72), 3.68 (95% CI: 1.80–7.50), and 3.16 (95% CI: 1.67–5.99) times’ increase in the odds of STI positivity for the respective sexual activities (Figure 4). Nevertheless, our analysis did not suggest a statistically significant association between STI positivity and barrier protection usage frequency (Supplementary Figure 7).

**Figure 4:**
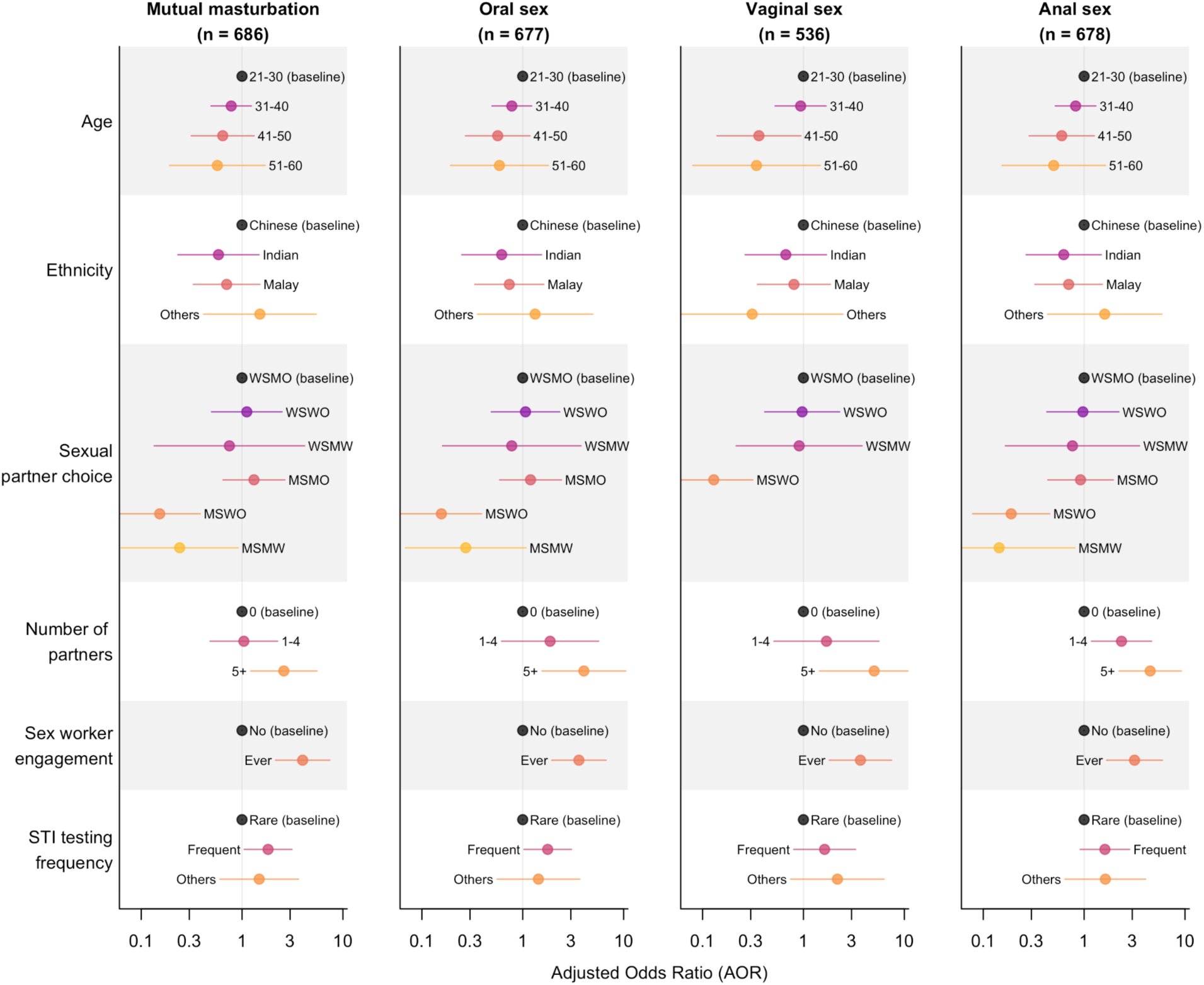
Adjusted odds ratio of STI positivity status for socio-demographics, partnership-based sexual orientation, number of sexual partners in the past year, engagement with sex workers, and STI testing frequency, stratified by sexual activity type (columns). These include point estimates (dots) and corresponding 95% CIs (lines). Extreme estimates due to insufficient samples (e.g., vaginal sex among MSMO) are not shown. Individuals who had never tested for STIs were excluded from the analysis. Rare STI testing is defined as testing once a year and frequent testing refers to testing once every three or six months. Please also note the x-axes are presented on a logarithmic scale.

## Discussion

This study provides insights into the diversity and distribution of sexual behaviours across the multi-ethnic Singapore population. By capturing detailed behavioural patterns, including partner numbers, types of sexual activity, and engagement with sex workers, across both heterosexual and non-heterosexual populations, the study enhances understanding of sexual behaviours that have been underrepresented in regional studies. In particular, the fitted distributions for the number of sexual partners across different sexual activities, alongside reported frequencies of participation in these activities, offer a more granular characterisation of sexual contact patterns than previously understood. Such localised data offer behavioural insights into transmission risk across sociodemographic groups and can better inform the parameterisation of STI models, enabling them to more accurately reflect behavioural patterns and population heterogeneity in Singapore.

Due to the lack of contemporary data in Singapore and many other Asian countries, STI modelling studies in Singapore have used partner number distributions and sexual activity assumptions drawn from Western contexts^25^. However, the number of partners and the participation in sexual practices likely vary between western countries and countries in Asia. This is corroborated by higher participation in partnered sexual activities in the past year reported by the NATSAL-3 survey in Britain with 82.1% of men and 77.7% of women between 16 – 74 years old who had at least one opposite sex partner over the past year^26^. In contrast, our results indicated that the participation in any type of sexual activity (mutual masturbation, oral sex, vaginal sex, and anal sex) in the past year was lower in Singapore than in Britain, at 71.7% (95% CI: 68.7% - 74.6%) and 56.6% (95% CI: 52.7% - 60.6%) of male and female participants aged 21 – 60 years old. This highlights the need for localised data when modelling STIs in the region. By providing behavioural estimates in Singapore, this study lays essential groundwork for more contextually accurate STI transmission models and more targeted public health interventions.

Marked sex- and partnership orientation-based differences in reported sexual activity were observed, particularly among individuals with opposite-sex partners. Male participants were more likely than female participants to report recent sexual activity across all four sexual activities (mutual masturbation, oral sex, vaginal sex, anal sex) and reported higher frequencies of engaging in these acts with female partners, than female respondents did with male partners. This asymmetry could reflect actual differences in behaviours, but may be influenced by gendered reporting biases, such as social desirability or cultural stigma, that differentially affect willingness to disclose sexual activity, especially among women^27,28^. In addition, WSW with at least one sexual partner reported lower frequencies of engaging in mutual masturbation, oral sex, and vaginal sex across different recall periods (Fig. 1). However, among those who were sexually active, a notable proportion reported high partner numbers (>5 partners in the past year). A similar pattern was observed among MSM, suggesting substantial variation within these groups: while many report low or moderate sexual activity, a smaller subset is more sexually active.

Engagement with sex workers was significantly more common among males than females, with males, across all partnership-based sexual orientation categories, having higher odds of engaging with sex workers within each of the regression models stratified by the four sexual activities (mutual masturbation, oral sex, vaginal sex, anal sex). While fewer females reported such engagement overall, variations by partnership-based sexual orientation were still apparent, with WSW reporting higher rates of engagement with sex workers than women who have sex exclusively with men (WSMO). These observations challenge prevailing assumptions in Singapore, where research on sex worker clientele has primarily focused on heterosexual men^12,13^, and point to the need for broader recognition of behavioural diversity. Importantly, individuals reporting five or more sexual partners in the past year were significantly more likely to have engaged with sex workers across all sexual activity types. This co-occurrence of different sexual behaviours, such as transactional sex, partner concurrency, and high partner numbers, has been documented in various settings globally, including among male factory workers in China^29^, substance-using youth in the United States^30^, and adults in rural communities in Kenya and Uganda^31^. Recognising co-occurrence of different sexual behaviours is important for both surveillance and intervention design. Surveillance strategies that analyse each risk factor independently may underestimate the threat posed by individuals with co-occurring behaviours, and interventions targeting one behaviour at a time may miss those most likely to benefit. Instead, using combined indicators such as partner number and partner type (sex worker or not) can enhance identification of high-transmission subgroups and guide more efficient intervention efforts like targeted STI screening, prevention outreach, and linkage to care.

Among individuals who reported ever testing for STIs, those who tested more frequently had higher self-reported positivity. This could reflect risk-aware behaviour, with higher-risk individuals proactively seeking testing, but could also indicate reverse causality, where symptomatic individuals or those previously diagnosed may test more often. This is important for interpreting surveillance data, as higher positivity rates among frequent testers may represent greater detection rather than elevated risk alone. Additionally, higher levels of sexual activity and reported engagement with sex workers were both associated with greater odds of self-reported STI positivity. While these associations align with existing understandings of STI risk, reverse causality remains a possibility, where individuals who test positive may retrospectively report more sexual activity or more carefully recall engagements with sex workers. This underscores the importance of targeting sexual health interventions and counselling toward subpopulations indicating these behaviours, to efficiently reduce transmission risks.

Interestingly, we did not find a significant association between self-reported STI positivity, and the frequency of barrier protection use. This finding may indicate measurement biases, including potential social desirability bias in reporting sensitive behaviours such as barrier protection usage and STI diagnoses. The relatively high non-response rates for questions on both barrier protection use and STI positivity suggest some participants may have felt uncomfortable disclosing these sensitive details, leading to reporting bias (Supplementary Table 3, Supplementary Figure 7). Given these limitations, the absence of a clear association should be interpreted with caution. Nevertheless, it highlights the importance of complementing barrier protection education with comprehensive behavioural risk-reduction strategies, including regular STI screening and robust partner notification programs, rather than relying on protective behaviours.

Several limitations should be acknowledged when interpreting our findings. First, our results cannot be generalized to older individuals (aged 61+), as they constituted only 4.4% of our sample, coupled with notably high non-response rates regarding specific sexual activity in this group. Therefore, this age group was removed from further analysis, restricting our ability to reliably infer sexual behaviour patterns within older populations. Second, despite our use of anonymous online surveys, social desirability bias may still have influenced responses, potentially leading to under-reporting of behaviours viewed as socially undesirable or stigmatized. Consequently, actual prevalence rates of higher-risk sexual activities may be underestimated. Thirdly, employing specific web communities and online survey methods could have introduced sampling bias, excluding less technologically inclined individuals and thus, potentially impacting the representativeness of our findings. Finally, to ensure sufficient sample sizes for characterising sexual behaviours among sexual minority groups, we actively recruited individuals identifying as MSM and WSW through LGBTQ-focused platforms. While this approach improved representation within these subgroups, the overall sample is not representative of the general population by sexual orientation. Accordingly, findings should be interpreted as reflective within, rather than across, each partnership-based sexual orientation category.

This study advances our understanding of contemporary sexual behaviours in Singapore, providing critical insights into diverse sexual practices and patterns relevant for STI prevention and modelling. By identifying nuanced differences based on sex, sexual orientation, and risk behaviours, the findings facilitate targeted and contextually appropriate public health interventions. Future research should aim to address identified limitations by engaging broader demographic groups, particularly older adults and less digitally accessible populations, ensuring more comprehensive representation. Continued surveillance, paired with interventions tailored to behavioural subgroups exhibiting elevated risk, will be essential in effectively mitigating STI transmission in Singapore.

## Data Availability

The data that support the findings of this study are not publicly available due to ethical/privacy restrictions but are available from the corresponding author (BD) on reasonable request.

## Author Contributions

All authors contributed to the conceptualisation and design of the study. JA, GG, RT, and BD coordinated data collection. SJ and JA led the methodology and data analysis. JA, SJ, and BD drafted the initial manuscript. All authors revised and edited the draft and approved the final version. BD provided supervision throughout the study and is the guarantor.

## Patient and Public Involvement

The public were not involved in the design, or conduct, or reporting, or dissemination plans of our research.

## References

1. Liu H, Waite L, Shen S, Wang D. Is Sex Good for Your Health? A National Study on Partnered Sexuality and Cardiovascular Risk Among Older Men and Women. J Health Soc Behav. 2016 Sep;57(3):276.

2. Mollaioli D, Sansone A, Ciocca G, Limoncin E, Colonnello E, Di Lorenzo G, et al. Benefits of Sexual Activity on Psychological, Relational, and Sexual Health During the COVID-19 Breakout. J Sex Med. 2020 Oct 23;18(1):35–49.

3. Hogben M, Leichliter J, Aral SO. An Overview of Social and Behavioral Determinants of STI. In: Cristaudo A, Giuliani M, editors. Sexually Transmitted Infections : Advances in Understanding and Management [Internet]. Cham: Springer International Publishing; 2020 [cited 2024 Oct 7]. p. 25–45. Available from: 10.1007/978-3-030-02200-6_3

4. United Nations Population Fund, United Nations Educational, Scientific and Cultural Organization. Sexual and reproductive health of young people in Asia and the Pacific [Internet]. 2015. Available from: https://asiapacific.unfpa.org/sites/default/files/pub-pdf/UNFPA%20SHR%20YP%20AP_2015%20for%20web-final.pdf

5. Junxuan Y, Zakarya Z, Ja’afar S, Jusoff K, Idris N. Exploring taboo culture: a cross-cultural analysis of taboos in China and Malaysia. Cogent Arts Humanit. 2025 Dec 31;12(1):2442828.

6. Wellings K, Collumbien M, Slaymaker E, Singh S, Hodges Z, Patel D, et al. Sexual behaviour in context: a global perspective. The Lancet. 2006 Nov 11;368(9548):1706–28.

7. Tan RKJ, Chan YY, Bin Ibrahim MA, Ho LP, Lim OZ, Choong BCH, et al. Potential interactions between the pathways to diagnosis of HIV and other STIs and HIV self-testing: insights from a qualitative study of gay, bisexual and other men who have sex with men in Singapore. Sex Transm Infect. 2021 May;97(3):215–20.

8. Tan RKJ, O’Hara CA, Koh WL, Le D, Tan A, Tyler A, et al. Delineating patterns of sexualized substance use and its association with sexual and mental health outcomes among young gay, bisexual and other men who have sex with men in Singapore: a latent class analysis. BMC Public Health. 2021 May 31;21(1):1026.

9. Tan RKJ, Ho V, Sherqueshaa S, Dee W, Lim JM, Lo JJM, et al. The Impact of the Coronavirus Disease (COVID-19) on the Health and Social Needs of Sex Workers in Singapore. Arch Sex Behav. 2021 Jul;50(5):2017–29.

10. Wong ML, Chan RKW, Chua WL, Wee S. Sexually Transmitted Diseases and Condom Use Among Female Freelance and Brothel-Based Sex Workers in Singapore: Sex Transm Dis. 1999 Nov;26(10):593–600.

11. Wong ML, Chan R, Tan HH, Yong E, Lee L, Cutter J, et al. Sex work and risky sexual behaviors among foreign entertainment workers in urban Singapore: findings from Mystery Client Survey. J Urban Health Bull N Y Acad Med. 2012 Dec;89(6):1031–44.

12. Lim RBT, Wong ML, Tan PH, Govender M. Heterosexual men who patronise entertainment establishments versus brothels in an Asian urban setting – which group practises riskier sexual behaviours? BMC Public Health. 2015 Aug 14;15:777.

13. Wong ML, Koh TT, Tjahjadi S, Govender M. Men seeking sex online practise riskier sexual behaviours than men frequenting brothels: survey findings from Singapore. Sex Transm Infect. 2014 Aug;90(5):401–7.

14. Lim RBT, Wong ML, Cook AR, Brun C, Chan RKW, Sen P, et al. Determinants of Chlamydia, Gonorrhea, and Coinfection in Heterosexual Adolescents Attending the National Public Sexually Transmitted Infection Clinic in Singapore. Sex Transm Dis. 2015 Aug;42(8):450–6.

15. Ng JYS, Chan RKW, Chio MTW, Lim RBT, Koh D, Wong ML. An Abstinence and Safer Sex Intervention for Adolescents Attending the Public Sexually Transmitted Infection Clinic in Singapore. J Adolesc Health. 2018 Jun;62(6):737–46.

16. Heng BH, Lee HP, Kok LP, Ong YW, Ho ML. A survey of sexual behaviour of Singaporeans. Ann Acad Med Singapore. 1992 Nov;21(6):723–9.

17. Tan PL. Stress, Fatigue, and Sexual Spontaneity Among Married Couples in a High-Stress Society: Evidence from Sex Diary Data from Singapore. Arch Sex Behav. 2021 Aug;50(6):2579–88.

18. Tan PL. Changes in Frequency and Patterns of Marital Sexual Activity During COVID-19: Evidence From Longitudinal Data Prior to, During and After Lockdown in Singapore. J Sex Med. 2022 Feb;19(2):188–200.

19. Tan RKJ, O’Hara CA, Kumar N. Partnership status, living arrangements, and changes in sexual behaviour and satisfaction during the COVID-19 lockdown: insights from an observational, cross-sectional online survey in Singapore. Sex Health. 2021 Nov;18(5):366–77.

20. Caltabiano M, Castiglioni M, De-Rose A. Changes in the sexual behaviour of young people: introduction. Genus. 2020 Dec;76(1):38, s41118-020-00107–1.

21. Lewis R, Tanton C, Mercer CH, Mitchell KR, Palmer M, Macdowall W, et al. Heterosexual Practices Among Young People in Britain: Evidence From Three National Surveys of Sexual Attitudes and Lifestyles. J Adolesc Health. 2017 Dec 1;61(6):694– 702.

22. Lumley T. survey: analysis of complex survey samples [Internet]. 2024. Available from: http://r-survey.r-forge.r-project.org/survey/

23. Department of Statistics, Singapore. Population and Population Structure [Internet]. 2025 [cited 2025 Mar 24]. Available from: https://www.singstat.gov.sg/find-data/search-by-theme/population/population-and-population-structure/latest-data

24. R Core Team. R: A Language and Environment for Statistical Computing [Internet]. Vienna, Austria: R Foundation for Statistical Computing; 2023. Available from: https://www.R-project.org/

25. Gan G, Janhavi A, Tong G, Lim JT, Dickens BL. The need for pre-emptive control strategies for mpox in Asia and Oceania. Infect Dis Model. 2024 Mar 1;9(1):214–23.

26. Mercer CH, Tanton C, Prah P, Erens B, Sonnenberg P, Clifton S, et al. Changes in sexual attitudes and lifestyles in Britain through the life course and over time: findings from the National Surveys of Sexual Attitudes and Lifestyles (Natsal). The Lancet. 2013 Nov;382(9907):1781–94.

27. Clark S, Kabiru C, Zulu E. Do Men and Women Report Their Sexual Partnerships Differently? Evidence from Kisumu, Kenya. Int Perspect Sex Reprod Health. 2011 Dec;37(4):181–90.

28. King BM. The Influence of Social Desirability on Sexual Behavior Surveys: A Review. Arch Sex Behav. 2022 Apr 1;51(3):1495–501.

29. Zhang K, Chen S, Zhu S, Fang Y, Zou H, Cai Y, et al. Multifaceted Determinants of Sexual Intercourse with Non-Regular Female Sex Partners and Female Sex Workers among Male Factory Workers in China—A Cross-Sectional Survey. Int J Environ Res Public Health. 2022 Jan;19(23):16008.

30. Patton RA, Cunningham RM, Blow FC, Zimmerman MA, Booth BM, Walton MA. Transactional Sex Involvement: Exploring Risk and Promotive Factors Among Substance-Using Youth in an Urban Emergency Department. J Stud Alcohol Drugs. 2014 Jul;75(4):573–9.

31. Okiring J, Getahun M, Gutin SA, Lebu S, Lee J, Maeri I, et al. Sexual partnership concurrency and age disparities associated with sexually transmitted infection and risk behavior in rural communities in Kenya and Uganda. Int J Infect Dis. 2022 Jul 1;120:158–67.

