## Supplementary Information for "Partnership-based sexual behaviours among adults in Singapore"

### Questionnaire

1. Survey with General Population
   (As part of the project on Queer Men in Singapore: Digital Intimacies, Sexual Behaviors, and Monkeypox Perceptions)

   Please read the following document: Participant information sheet (PIS)

   ***Instructions:*** Please check off all of the following as you read them. You can only proceed to the survey when you do so.

   ▢ I have read and understood the Participant Information Sheet (PIS).
   ▢ I understand that my participation in this study is voluntary and anonymous.
   ▢ I understand that I can choose to pause to take a break from, return to, stop the survey, or skip or prefer not to answer any question for any reason, with no consequence.
   ▢ I understand that under Section 23(1) of the Infectious Diseases Act, persons living with HIV are required to inform their sexual partners of the risk of HIV transmission, so that their sexual partners can make an informed decision whether to proceed with the sexual encounter, and to take the necessary precautions to protect themselves, which will in turn reduce the risk of HIV transmission. A person convicted of an offence under the Infectious Diseases Act and its Regulations is liable to a fine of up to $10,000, or up to six months' imprisonment, or both.
   ▢ I understand that if I have any questions about the Participant Information Sheet (PIS) or the study, I can reach out to the researchers (whose contact information is given in the PIS) at any time before, after, and while doing this online survey.
   ▢ I declare that I am at least 21 years old or older and have resided in Singapore for at least the past 6 months.
   ▢ I consent to take part in this research study.

2. Before you complete the survey, please complete the captcha below
3. What is your age?
   - Please specify below ______________________________
   - Prefer not to say
4. What is your sex assigned at birth?
   - Male
   - Female
   - Others (please specify below): ______________________________
   - Prefer not to say (Skip to end of the survey after Q 12)
5. What is your gender identity?
   - Man
   - Woman
   - Others (please specify below): ______________________________
   - Prefer not to say (Skip to end of the survey after Q 12)
6. What is your ethnicity?
   - Chinese
   - Malay
   - Indian
   - Others (please specify below): ______________________________
   - Prefer not to say
7. What is your legal relationship status?
   - Single
   - Married
   - Separated
   - Divorced
   - Other (please specify below): ______________________________
   - Prefer not to say

8. What is your resident status?
   - o Singapore citizen
   - o Permanent resident
   - o Work pass holder (e.g., Work permit, S-Pass, Employment Pass)
   - o Other long-term resident passes (e.g., LTVP)
   - o Others (please specify below) ____________________________________________
   - o Prefer not to say
9. What is your religion?
   - o Buddhism
   - o Taoism
   - o Christianity
   - o Islam
   - o Hinduism
   - o Other religion (please specify below):
     ________________________________________
   - o Prefer not to say
10. What is your gross monthly income (before tax and CPF reductions)?
    - o Not earning an income
    - o <SGD2000
    - o SGD2000 - SGD4999
    - o SGD5000 - SGD7999
    - o SGD8000 - SGD9999
    - o SGD10000 and above
    - o Prefer not to say
11. What is your sexual orientation?
    - o Heterosexual/Straight
    - o Homosexual/Gay/Lesbian
    - o Bisexual
    - o Queer
    - o Questioning
    - o Others (please specify below): ____________________________________________
    - o Prefer not to say
12. What is/are the biological sex of previous & current sexual partners?
    - o Male
    - o Female
    - o Both
    - o No current or previous sexual partners (Skip to end of survey)
    - o Prefer not to say (Skip to end of survey)
13. Have you engaged with a sex worker? (*We define sex workers as individuals who receive money or goods in exchange for sexual services.)*
    - o No (Skip to Question 16)
    - o Yes
    - o Prefer not to say (Skip to Question 16)
14. If yes, what is the gender of the sex worker?
    - o Male
    - o Female
    - o Both
    - o Others
    - o Prefer not to say
15. (Please enter number for all boxes to move on. If you prefer not to answer, enter NIL for all the boxes) How many sex workers
    - o Have you ever been with? ____________________________________________
    - o Have you been with in the past 1 year? ____________________________________
    - o Have you been with in the past 6 months? __________________________________
    - o Have you been with in the past 3 months? __________________________________
16. Do you use sex toys? (*Sex toys are objects that are used for sexual stimulation which come into contact with genitals)*
    - o No (Skip to next relevant section)
    - o Yes
    - o Prefer not to say (Skip to next relevant section)

17. Do you use the same sex toy on multiple sexual partners?
    - No (1)
    - Yes (2)
    - Prefer not to say (3)

Participants answered the following sections based on their sex assigned at birth (Question 4) and biological sex of their sexual partners (Question 12).

**Section 1:** ***For all who have had a female partner (Question 12 = Female/Both)***
**Mutual masturbation with a female partner**

18. Have you participated in mutual masturbation *(using hands or toys to stimulate each other's genitals)* with a female partner?
    - No (Skip Question 19 & 20)
    - Yes
    - Prefer not to say (Skip Question 19 & 20)
19. (Please enter number for all boxes to move on. If you prefer not to answer, enter NIL for all the boxes) What is the number of female partners you have engaged in mutual masturbation with
    - In the past 1 year?________________________________
    - In the past 6 months? ________________________________
    - In the past 3 months? ________________________________
20. (Please enter number for all boxes to move on. If you prefer not to answer, enter NIL for all the boxes) How many times have you engaged in mutual masturbation with a female partner
    - In the past 1 year?________________________________
    - In the past 6 months? ________________________________
    - In the past 3 months?________________________________

**Section 2:** ***For all who have had a female partner (Question 12 = Female/Both)***
**Oral sex with a female partner**

21. Have you performed or received oral sex with a female partner?
    - No (Skip Questions 22 - 25)
    - Yes
    - Prefer not to say (Skip Questions 22 - 25)
22. What is your preferred action?
    - Receiving
    - Performing
    - Both
    - Prefer not to say
23. (Please enter number for all boxes to move on. If you prefer not to answer, enter NIL for all the boxes) What is the number of female partners you have performed or received oral sex with
    - In the past 1 year?________________________________
    - In the past 6 months? ________________________________
    - In the past 3 months?________________________________
24. (Please enter number for all boxes to move on. If you prefer not to answer, enter NIL for all the boxes) How many times have you performed or received oral sex with a female partner
    - In the past 1 year?________________________________
    - In the past 6 months? ________________________________
    - In the past 3 months?________________________________
25. How often is test protection used (condoms/dental dams) when performing/receiving oral sex?
    - All the time
    - At least half the time
    - Occasionally
    - Never
    - Prefer not to say

**Section 3A:** ***For males (Q4) with female partner(s) (Question 12 = Female/Both)***
**Vaginal sex with a female partner**

26. Have you had vaginal sex with a female partner?
    - No (Skip Questions 27 - 29)
    - Yes
    - Prefer not to say (Skip Questions 27 - 29)

27. (Please enter number for all boxes to move on. If you prefer not to answer, enter NIL for all the boxes) What is the number of female partners you have had vaginal sex with
    - In the past 1 year?______________________________________________
    - In the past 6 months? ______________________________________________
    - In the past 3 months?______________________________________________

28. (Please enter number for all boxes to move on. If you prefer not to answer, enter NIL for all the boxes) How many times have you had vaginal sex with a female partner
    - In the past 1 year?______________________________________________
    - In the past 6 months? ______________________________________________
    - In the past 3 months?______________________________________________
29. How often is barrier protection used (condoms) when having vaginal sex?
    - All the time
    - At least half the time
    - Occasionally
    - Never
    - Prefer not to say

**Section 4A:** ***For males (Q4) with female partner(s) (Question 12 = Female/Both)***
**Anal sex with a female partner**

30. Have you had anal sex with a female partner?
    - No (Skip Questions 31 - 33)
    - Yes
    - Prefer not to say (Skip Questions 31 - 33)
31. (Please enter number for all boxes to move on. If you prefer not to answer, enter NIL for all the boxes) What is the number of female partners you have had anal sex with
    - In the past 1 year?______________________________________________
    - In the past 6 months? ______________________________________________
    - In the past 3 months?______________________________________________
32. (Please enter number for all boxes to move on. If you prefer not to answer, enter NIL for all the boxes) How many times have you had anal sex with a female partner
    - In the past 1 year?______________________________________________
    - In the past 6 months? ______________________________________________
    - In the past 3 months?______________________________________________
33. How often is barrier protection used (condoms) when having anal sex?
    - All the time
    - At least half the time
    - Occasionally
    - Never
    - Prefer not to say

**Section 3B:** ***For females (Q4) with female partner(s) (Question 12 = Female/Both)***
**Vaginal sex with a female partner**

34. Have you had vaginal sex (using insertive sex toys) with a female partner?
    - No (Skip Questions 35 - 37)
    - Yes
    - Prefer not to say (Skip Questions 35 - 37)
35. (Please enter number for all boxes to move on. If you prefer not to answer, enter NIL for all the boxes) What is the number of female partners you have had vaginal sex (using insertive sex toys) with
    - In the past 1 year?______________________________________________
    - In the past 6 months? ______________________________________________
    - In the past 3 months?______________________________________________
36. (Please enter number for all boxes to move on. If you prefer not to answer, enter NIL for all the boxes) How many times have you had vaginal sex (using insertive sex toys) with a female partner
    - In the past 1 year?______________________________________________
    - In the past 6 months? ______________________________________________
    - In the past 3 months?______________________________________________

37. How often is barrier protection used (external condoms with insertive sex toys) when having vaginal sex?
    - o All the time
    - o At least half the time
    - o Occasionally
    - o Never
    - o Prefer not to say

**Section 4B:** ***For females (Q4) with female partner(s) (Question 12 = Female/Both)***
**Anal sex with a female partner**

38. Have you had anal sex (using insertive sex toys) with a female partner?
    - o No (Skip Questions 39 - 41)
    - o Yes
    - o Prefer not to say (Skip Questions 39 - 41)
39. (Please enter number for all boxes to move on. If you prefer not to answer, enter NIL for all the boxes) What is the number of female partners you have had anal sex (using insertive sex toys) with
    - o In the past 1 year?____________________
    - o In the past 6 months? ____________________
    - o In the past 3 months?____________________
40. (Please enter number for all boxes to move on. If you prefer not to answer, enter NIL for all the boxes) How many times have you had anal sex (using insertive sex toys) with a female partner
    - o In the past 1 year?____________________
    - o In the past 6 months? ____________________
    - o In the past 3 months?____________________
41. How often is barrier protection used (external condoms with insertive sex toys) when having anal sex?
    - o All the time
    - o At least half the time
    - o Occasionally
    - o Never
    - o Prefer not to say

**Section 3C:** ***For (Q4 = others) with female partner(s) (Question 12 = Female/Both)***
**Vaginal sex with a female partner**

42. Have you had vaginal sex (including the use of insertive sex toys) with a female partner?
    - o No (Skip Questions 43 - 45)
    - o Yes
    - o Prefer not to say (Skip Questions 43 - 45)
43. (Please enter number for all boxes to move on. If you prefer not to answer, enter NIL for all the boxes) What is the number of female partners you have had vaginal sex (including the use of insertive sex toys) with
    - o In the past 1 year?____________________
    - o In the past 6 months? ____________________
    - o In the past 3 months?____________________
44. (Please enter number for all boxes to move on. If you prefer not to answer, enter NIL for all the boxes) How many times have you had vaginal sex (including the use of insertive sex toys) with a female partner
    - o In the past 1 year?____________________
    - o In the past 6 months? ____________________
    - o In the past 3 months?____________________

45. How often is barrier protection used (including external condoms with insertive sex toys) when having vaginal sex?
    - o All the time
    - o At least half the time
    - o Occasionally
    - o Never
    - o Prefer not to say

**Section 4B:** ***For (Q4 = others) with female partner(s) (Question 12 = Female/Both)***
**Anal sex with a female partner**

46. Have you had anal sex (including the use of insertive sex toys) with a female partner?
    - No (Skip Questions 47 - 49)
    - Yes
    - Prefer not to say (Skip Questions 47 - 49)
47. (Please enter number for all boxes to move on. If you prefer not to answer, enter NIL for all the boxes) What is the number of female partners you have had anal sex (including the use of insertive sex toys) with
    - In the past 1 year?__________________________
    - In the past 6 months? __________________________
    - In the past 3 months?__________________________
48. (Please enter number for all boxes to move on. If you prefer not to answer, enter NIL for all the boxes) How many times have you had anal sex (including the use of insertive sex toys) with a female partner
    - In the past 1 year?__________________________
    - In the past 6 months? __________________________
    - In the past 3 months?__________________________
49. How often is barrier protection used (including external condoms with insertive sex toys) when having anal sex?
    - All the time
    - At least half the time
    - Occasionally
    - Never
    - Prefer not to say

**Section 5:** ***For all who have had a male partner (Question 12 = Male/Both)***
**Mutual masturbation with a male partner**

50. Have you participated in mutual masturbation *(using hands or toys to stimulate each other's genitals)* with a male partner?
    - No (Skip Question 51 & 52)
    - Yes
    - Prefer not to say (Skip Question 51 & 52)
51. (Please enter number for all boxes to move on. If you prefer not to answer, enter NIL for all the boxes) What is the number of male partners you have engaged in mutual masturbation with
    - In the past 1 year?__________________________
    - In the past 6 months? __________________________
    - In the past 3 months?__________________________
52. (Please enter number for all boxes to move on. If you prefer not to answer, enter NIL for all the boxes) How many times have you engaged in mutual masturbation with a male partner
    - In the past 1 year?__________________________
    - In the past 6 months? __________________________
    - In the past 3 months?__________________________

**Section 6:** ***For all who have had a male partner (Question 12 = Male/Both)***
**Oral sex with a male partner**

53. Have you performed or received oral sex with a male partner?
    - No (Skip Questions 54 - 57)
    - Yes
    - Prefer not to say (Skip Questions 54 - 57)
54. What is your preferred action?
    - Receiving
    - Performing
    - Both
    - Prefer not to say

55. (Please enter number for all boxes to move on. If you prefer not to answer, enter NIL for all the boxes) What is the number of male partners you have performed or received oral sex with
    - In the past 1 year?________________________________________________
    - In the past 6 months? ______________________________________________
    - In the past 3 months?______________________________________________
56. (Please enter number for all boxes to move on. If you prefer not to answer, enter NIL for all the boxes) How many times have you performed or received oral sex with a male partner
    - In the past 1 year?________________________________________________
    - In the past 6 months? ______________________________________________
    - In the past 3 months?______________________________________________
57. How often is barrier protection used (condoms/dental dams) when performing/receiving oral sex?
    - All the time
    - At least half the time
    - Occasionally
    - Never
    - Prefer not to say

**Section 7:** ***For non-males (Q4 = Female/ Others, OR Q5 = Woman/Others) with male partner(s) (Question 12 = Male/Both)***

**Vaginal sex with a male partner**

58. Have you had vaginal sex with a male partner?
    - No (Skip Questions 59 - 61)
    - Yes
    - Prefer not to say (Skip Questions 59 - 61)
59. (Please enter number for all boxes to move on. If you prefer not to answer, enter NIL for all the boxes) What is the number of male partners you have had vaginal sex with
    - In the past 1 year?________________________________________________
    - In the past 6 months? ______________________________________________
    - In the past 3 months?______________________________________________
60. (Please enter number for all boxes to move on. If you prefer not to answer, enter NIL for all the boxes) How many times have you had vaginal sex with a male partner
    - In the past 1 year?________________________________________________
    - In the past 6 months? ______________________________________________
    - In the past 3 months?______________________________________________
61. How often is barrier protection used (condoms) when having vaginal sex?
    - All the time
    - At least half the time
    - Occasionally
    - Never
    - Prefer not to say

**Section 8A:** ***For non-females (Q4 = Male/ Others) with male partner(s) (Question 12 = Male/Both)***

**Anal sex with a male partner**

62. Have you had anal sex with a male partner?
    - No (Skip Questions 63 - 66)
    - Yes
    - Prefer not to say (Skip Questions 63 - 66)
63. What is your preferred action?
    - Receiving
    - Performing
    - Both
    - Prefer not to say
64. (Please enter number for all boxes to move on. If you prefer not to answer, enter NIL for all the boxes) What is the number of male partners you have had anal sex with
    - In the past 1 year?________________________________________________
    - In the past 6 months? ______________________________________________
    - In the past 3 months?______________________________________________

65. (Please enter number for all boxes to move on. If you prefer not to answer, enter NIL for all the boxes) How many times have you had anal sex with a male partner
    - In the past 1 year?________________________________________
    - In the past 6 months? ________________________________________
    - In the past 3 months?________________________________________
66. How often is barrier protection used (condoms) when having anal sex?
    - All the time
    - At least half the time
    - Occasionally
    - Never
    - Prefer not to say

**Section 8A:** ***For females (Q4 = Female) with male partner(s) (Question 12 = Male/Both)***
**Anal sex with a male partner**

67. Have you had anal sex with a male partner?
    - No (Skip Questions 68 - 70)
    - Yes
    - Prefer not to say (Skip Questions 68 - 70)
68. (Please enter number for all boxes to move on. If you prefer not to answer, enter NIL for all the boxes) What is the number of male partners you have had anal sex with
    - In the past 1 year?________________________________________
    - In the past 6 months? ________________________________________
    - In the past 3 months?________________________________________
69. (Please enter number for all boxes to move on. If you prefer not to answer, enter NIL for all the boxes) How many times have you had anal sex with a male partner
    - In the past 1 year?________________________________________
    - In the past 6 months? ________________________________________
    - In the past 3 months?________________________________________
70. How often is barrier protection used (condoms) when having anal sex?
    - All the time
    - At least half the time
    - Occasionally
    - Never
    - Prefer not to say

All participants who had sexual partners were prompted to answer the following questions

71. Have you been diagnosed or tested positive for an STI before? This includes diagnosis for bacterial vaginosis or candidiasis (yeast infections).
    - No (Skip Questions 72 – 76)
    - Yes
    - Prefer not to say (Skip Questions 72 – 76)
72. Which STI(s) did you test positive for?
    - Bacterial Vaginosis
    - Candidiasis (yeast infection)
    - Chlamydia
    - Genital herpes
    - Genital warts
    - Gonorrhoea
    - Hepatitis B
    - Hepatitis C
    - HIV
    - Non-Gonococcal Urethritis
    - Syphilis
    - Others, Please specify: ________________________________________
    - Prefer not to say

73. What treatment did you receive for the STI?
    - o Please enter treatment below ______________________________
    - o Prefer not to say
74. Did you experience symptoms?
    - o Yes
    - o No (Skip Questions 75 & 76)
    - o Prefer not to say (Skip Questions 75 & 76)
75. How long did you wait before seeking treatment?
    - o Please enter length of time below: ______________________________
    - o Prefer not to say
76. Did you stop having sex after onset of symptoms?
    - o Yes
    - o No
    - o Prefer not to say
77. When was the last time you got an STI test?
    - o Within the last 3 months
    - o Within the last 6 months
    - o Within the last 1 year
    - o More than a year ago
    - o Never
    - o Prefer not to say

78. How regularly do you test for STIs?
    - o Never
    - o Once every 3 months
    - o Once every 6 months
    - o Once a year
    - o Others, please specify:______________________________
    - o Prefer not to say

**79. Reimbursement**

There is no reimbursement for participating in this survey.

However, you have the option to enter into a lucky draw for a chance to win $50 in Grab vouchers for your time. Your entry into the lucky draw is separate from the survey results.

Would you would like to enter into our lucky draw?
- o Yes (you will be redirected to another page)
- o No

### Collection of survey responses

#### Consent

Prior to the survey, participants were informed about the nature of questions asked and were provided the Participant Information Sheet (PIS), which they could download. Contact information of researchers involved in the study was provided in the PIS. Participants had to individually acknowledge points in the PIS to proceed with the survey. Participation was fully voluntary, with options to skip specific questions (Phases I and II), or prompts to click off the survey if they did not wish to disclose their information (Phase III).

#### Reimbursement

During Phase I and II, participants were offered to participate in a lucky draw to win SGD 50 in Grab vouchers (App for deliveries, mobility, and financial services) at the end of the survey. The form collecting participant information for the lucky draw was separate from the form that collected the survey responses to ensure that participants remained anonymous.

### Statistical analysis

*Distribution fitting*

Parameters for the Pareto distribution were estimated using bootstrapping. Specifically, we generated 100 bootstrapped samples from the raw data and the corresponding sample weights. We calculated the empirical cumulative distribution function, $F^{emp}(\cdot)$, for each bootstrapped dataset. We defined the loss function as

$$L(p) = \sum_i \left[\log\left(1 - F^{emp}(x_i)\right) - \log(1 - F_p(x_i))\right]^2 \cdot w_i.$$

In this, $\{x_i\}_i$ is a grid of all plausible values with a uniform step size $\delta = x_{i+1} - x_i$. $F_p(\cdot)$ is the cumulative density function for a Pareto distribution with parameters $p = \{n_0, \alpha\}$. To prevent bias toward sparse samples with high levels of sexual activity, we introduced a weight variable $w_i = 1_{i>1} \cdot (\log(x_i + 1) - \log(x_{i-1} + 1))^2 + 1_{i=1}$, where $1_y$ is a binary indicator function which equals 1 if and only if condition $y$ is satisfied and 0 otherwise. We then used the optim function in R to determine the parameter values, $\hat{p}$, which minimized loss function across all bootstrapped datasets.

*Logistic regression*

In the main analysis, three logistic regression models were considered:

i) Sexual partnership status ~ Age + Ethnicity + Partnership-based sexual orientation,
ii) Engagement with sex workers ~ Age + Ethnicity + Partnership-based sexual orientation + Number of sexual partners, and
iii) STI positivity ~ Age + Ethnicity + Partnership-based sexual orientation + Number of sexual partners + Engagement with sex workers + STI testing frequency.

Particularly, for model iii), we included only individuals who has ever tested for STIs, as those who had not undergone an STI test had unknown STI positivity status.

Regression analyses were also performed to investigate potential contributors to being highly sexually active or frequently engaging with sex workers. The following two models were constructed:

iv) High sexual activity level ~ Age + Ethnicity + Partnership-based sexual orientation, and
v) Frequent engagement with sex workers ~ Age + Ethnicity + Partnership-based sexual orientation + Number of sexual partners.

In addition, we examined the association between STI positivity and barrier protection usage frequency by adapting model iii) to:

vi) STI positivity ~ Age + Ethnicity + Partnership-based sexual orientation + Number of sexual partners + Engagement with sex workers + STI testing frequency + Barrier protection usage frequency.

Consistent with model iii), this model was applied to individuals who has ever tested for STI. This model was not applied to mutual masturbation as barrier protection is not relevant to this activity.

Across all models, sexual partnership status, number of sexual partners, and frequent engagement with sex workers, were assessed over the past year (i.e., using a one-year recall period). These variables, together with engagement with sex workers, were defined separately for each of the four sexual activities (mutual masturbation, oral sex, vaginal sex, and anal sex) unless otherwise specified. However, no specific timeframe was applied to socio-demographics (age and ethnicity in this context), partnership-based sexual orientation, engagement with sex workers, STI positivity, STI testing frequency, or barrier protection usage frequency. All variables included in the models were either binary or categorical, with their definitions and categories/possible values detailed in Table S1.

**Supplementary Table 1**. Regression model variables.

| Variable | Type | Definition | Categories/Possible values |
|---|---|---|---|
| Age | Categorical | Age group | • 21–30<br>• 31–40<br>• 41–50<br>• 51–60 |
| Ethnicity | Categorical | Ethnic group | ○ Chinese |

| | | | |
|---|---|---|---|
| | | | ○ Malay<br>○ Indian<br>○ Others |
| Partnership-based sexual orientation | Categorical | Sexual orientation determined by individual sexual behaviours, with sex defined as sex assigned at birth | • Men who have sex with men only (MSMO)<br>• Men who have sex with women only (MSWO)<br>• Men who have sex with both sexes (MSMW)<br>• Women who have sex with men only (WSMO)<br>• Women who have sex with women only (WSWO)<br>• Women who have sex with both sexes (WSMW)<br>• Others |
| Number of sexual partners | Categorical | Groups with varying sexual activity levels, classified based on the number of sexual partners in the past year | ○ 0<br>○ 1–4<br>○ 5+ |
| Sexual partnership status | Binary | Whether to have at least one sexual partner in the past year | • Yes (1)<br>• No (0) |
| Engagement with sex workers | Binary | Whether to have ever engaged with sex workers, who received money or goods in exchange for sexual services | ○ Ever (1)<br>○ No (0) |
| Frequent engagement with sex workers | Binary | Whether to have frequently engaged with sex workers in the past year | • Yes (1): individuals who engaged with five or more sex workers in the past year<br>• No (0) |
| STI positivity | Binary | Whether to have been diagnosed or tested positive for one or more STIs | ○ Yes (1)<br>○ No (0) |
| STI testing frequency | Categorical | How regularly an individual tests for STIs | • Never<br>• Rare: Once a year<br>• Frequent: Once every three or six months<br>• Others |
| Barrier protection usage frequency | Categorical | How frequent an individual uses barrier protection when having sex | • Never<br>• Occasionally<br>• 50%+: At least half the time<br>• Always: All the time<br>• Unknown |

### Additional analytical results

**Supplementary Table 2. Proportion (%) of missing responses for key survey questions among participants aged 21–60.**

| Question | 21–30 | 31–40 | 41–50 | 51–60 | Total |
|---|---|---|---|---|---|
| Mutual masturbation with males | 2.1 | 2.8 | 2.4 | 0.3 | 0.9 |
| Oral sex with males | 5.1 | 7.1 | 4.1 | 3.5 | 3.1 |
| Vaginal sex with males | 16.7 | 19.4 | 16 | 14.3 | 13 |
| Anal sex with males | 6.8 | 8 | 6 | 5.9 | 6.7 |
| Mutual masturbation with females | 1.7 | 1.4 | 1.9 | 1.3 | 3.1 |
| Oral sex with females | 3.7 | 3.1 | 3.3 | 3.8 | 7.6 |
| Vaginal sex with females | 4.7 | 4.5 | 3.7 | 4.3 | 9.9 |
| Anal sex with females | 5.1 | 4.4 | 4.2 | 5.4 | 10.3 |
| Engagement with sex workers | 13.9 | 19.5 | 9.4 | 10.0 | 14.3 |
| STI positivity status | 24.2 | 30.3 | 17.9 | 21.3 | 27.4 |

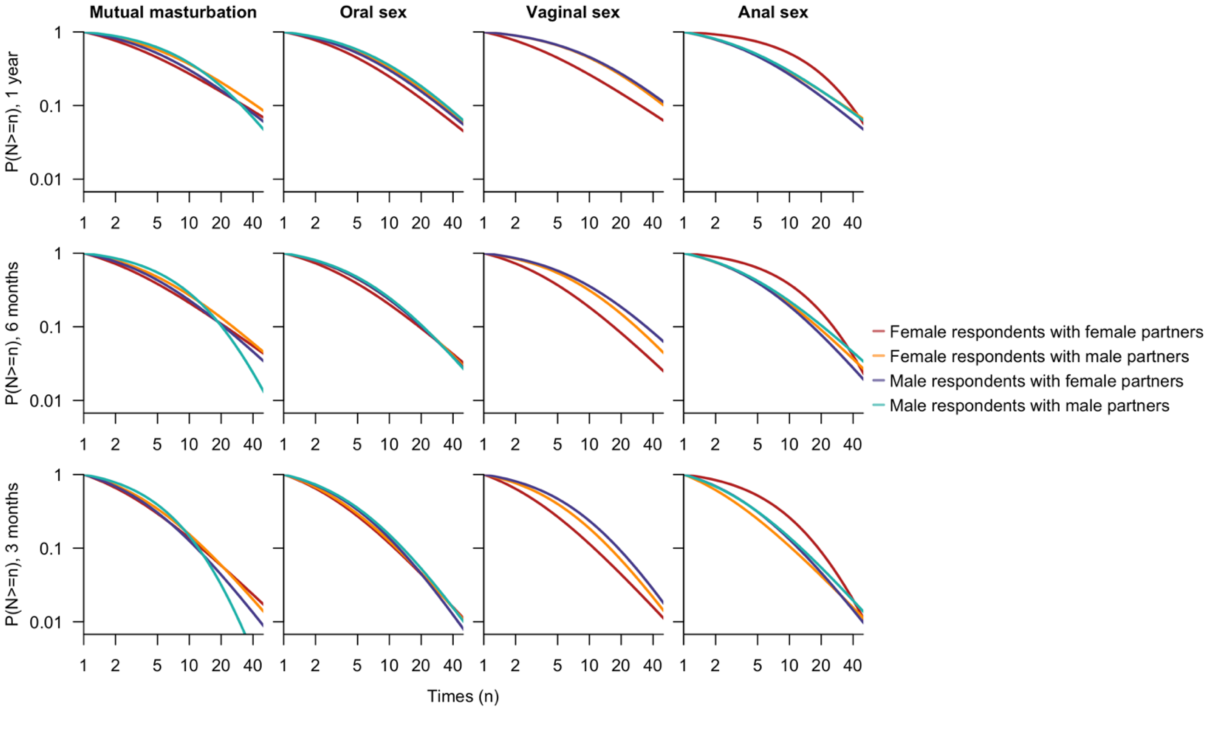

#### Supplementary Figure 1

Fitted distribution of the frequency of sexual activities (N) among individuals with sexual partners, stratified by the sex assigned at birth of survey respondents and their partners, type of sexual activity (columns), and recall period (rows). The distribution of the frequency of vaginal sex among males who have sex with males was not fitted or presented due to the small sample size. Both the x- and y-axes are displayed on a logarithmic scale.

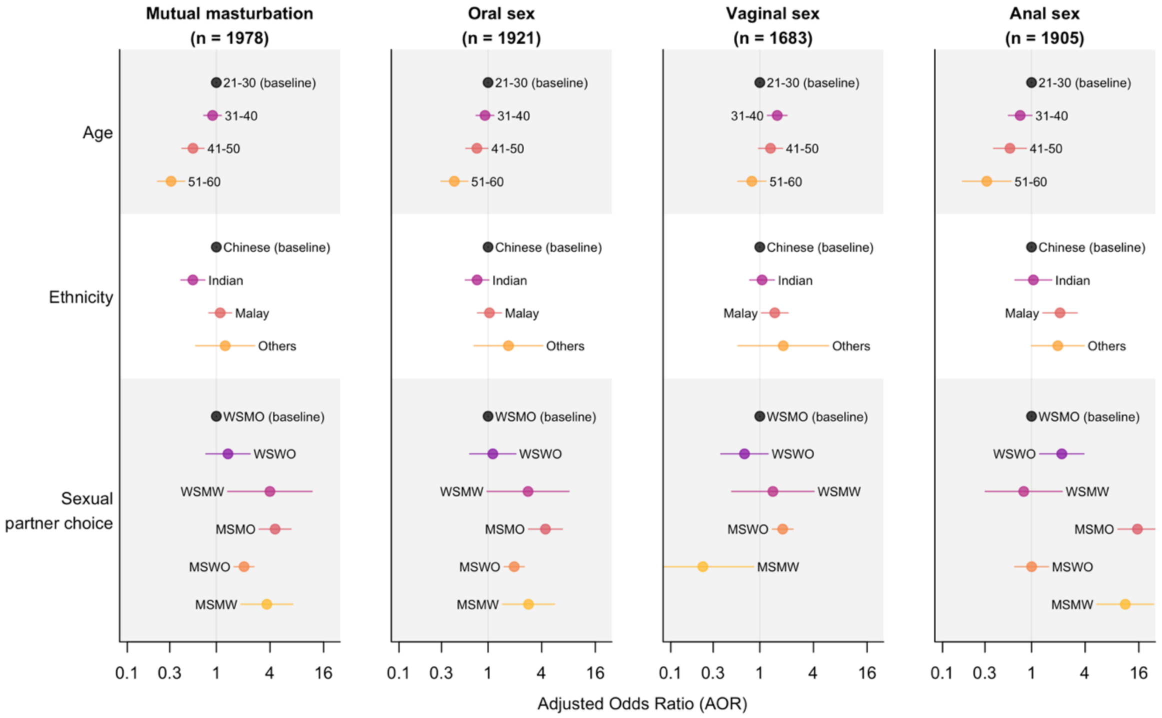

### Supplementary Figure 2

Adjusted odds ratio for socio-demographics and partnership-based sexual orientation defined by sex assigned at birth, stratified by sexual activity type (columns). These include point estimates (dots) and corresponding 95% Cis (lines), quantifying the factors' impacts on having at least one sexual partner in the past year. Extreme estimates due to insufficient samples (e.g., vaginal sex among MSMO) are not shown. Please also note the x-axes are presented on a logarithmic scale.

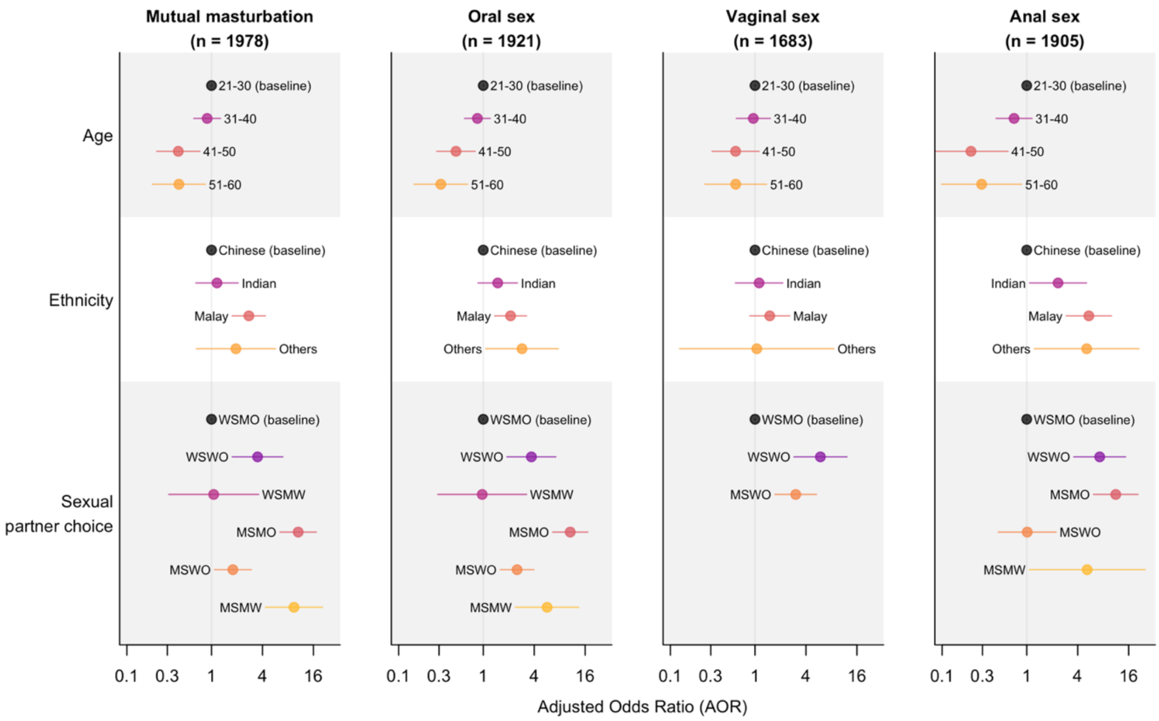

**Supplementary Figure 3**

Adjusted odds ratio for socio-demographics and partnership-based sexual orientation defined by sex assigned at birth, stratified by sexual activity type (columns). These include point estimates (dots) and corresponding 95% CIs (lines), quantifying the factors' impacts on being highly sexually active (i.e., having five sexual partners or more) in the past year. Extreme estimates due to insufficient samples (e.g., vaginal sex among MSMO) are not shown. Please also note the x-axes are presented on a logarithmic scale.

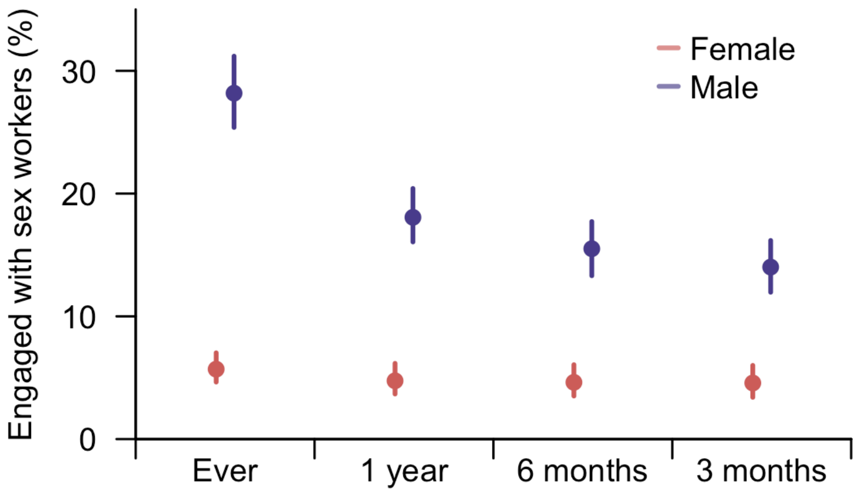

**Supplementary Figure 4**

Estimated proportion of individuals who reported having engaged with sex workers, stratified by sex assigned at birth and recall period. Dots represent estimated mean values and lines indicate the corresponding 95% CIs.

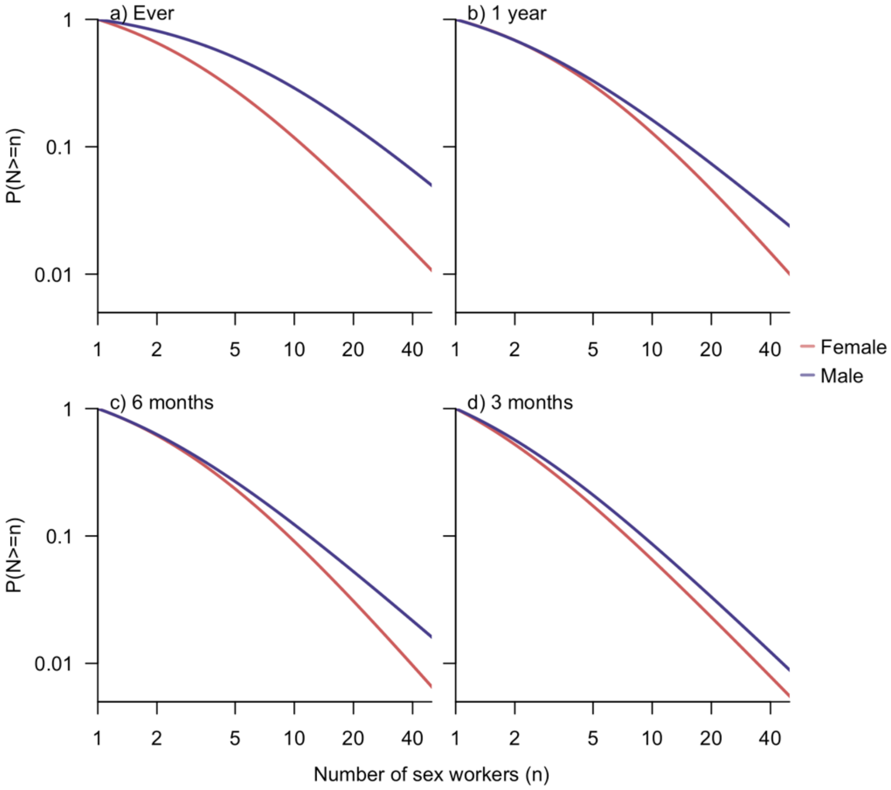

### Supplementary Figure 5

Fitted distribution of number of sex workers (N) an individual has engaged with among those who reported engaging with sex workers, stratified by sex assigned at birth and recall period. Both the x- and y-axes are displayed on a logarithmic scale.

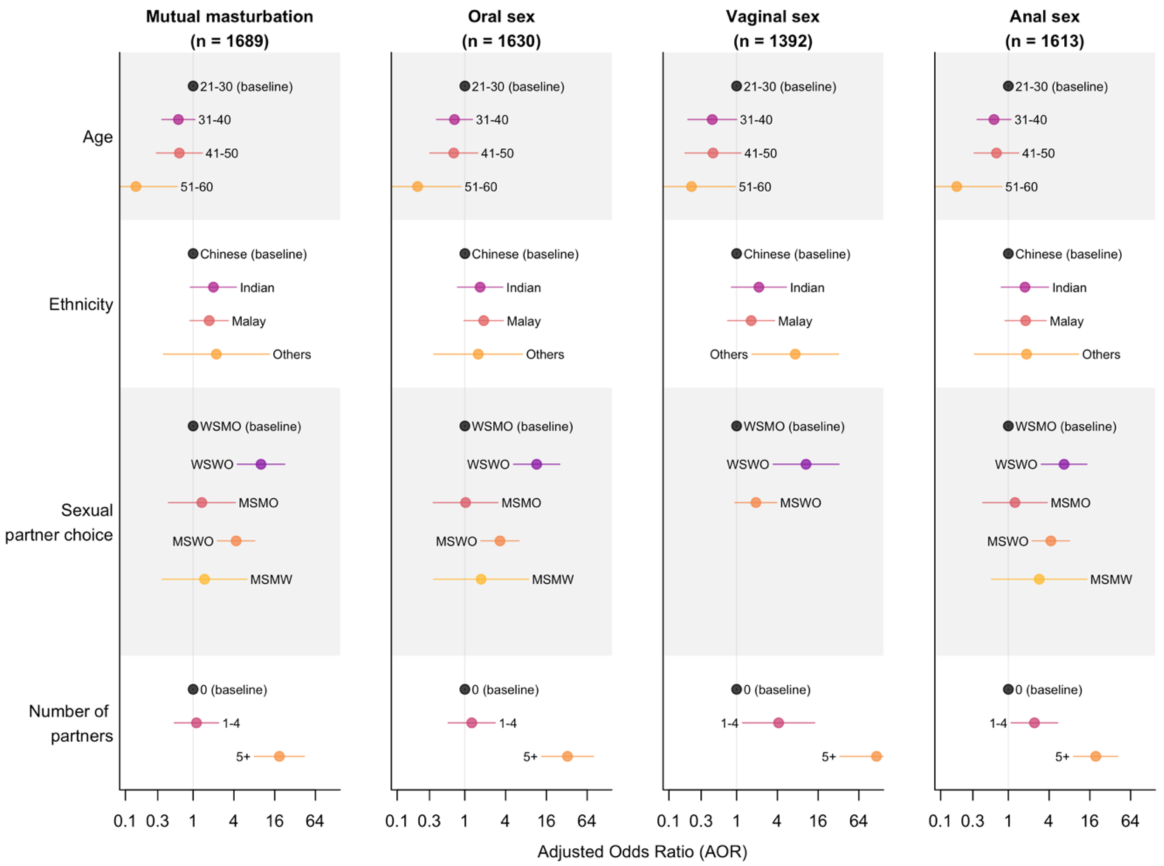

### Supplementary Figure 6

Adjusted odds ratio for socio-demographics, partnership-based sexual orientation defined by sex assigned at birth, and number of sexual partners in the past year, stratified by sexual activity type (columns). These include point estimates (dots) and corresponding 95% CIs (lines), quantifying the factors' impacts on frequent engagement with sex workers (i.e., having engaged with five or more sex workers in the past year). Extreme estimates due to insufficient samples (e.g., vaginal sex among MSMO) are not shown. Please also note the x-axes are presented on a logarithmic scale.

**Supplementary Table 3**. **STI positivity status among respondents aged 21–60, stratified by age group.**

| Age | Positive (n) | Negative (n) | Unknown (n) | Total (n) |
|---|---|---|---|---|
| 21–30 | 105 | 484 | 256 | 845 |
| 31–40 | 83 | 538 | 135 | 756 |
| 41–50 | 22 | 270 | 79 | 371 |
| 51–60 | 8 | 154 | 61 | 223 |
| Total | 218 | 1446 | 531 | 2195 |

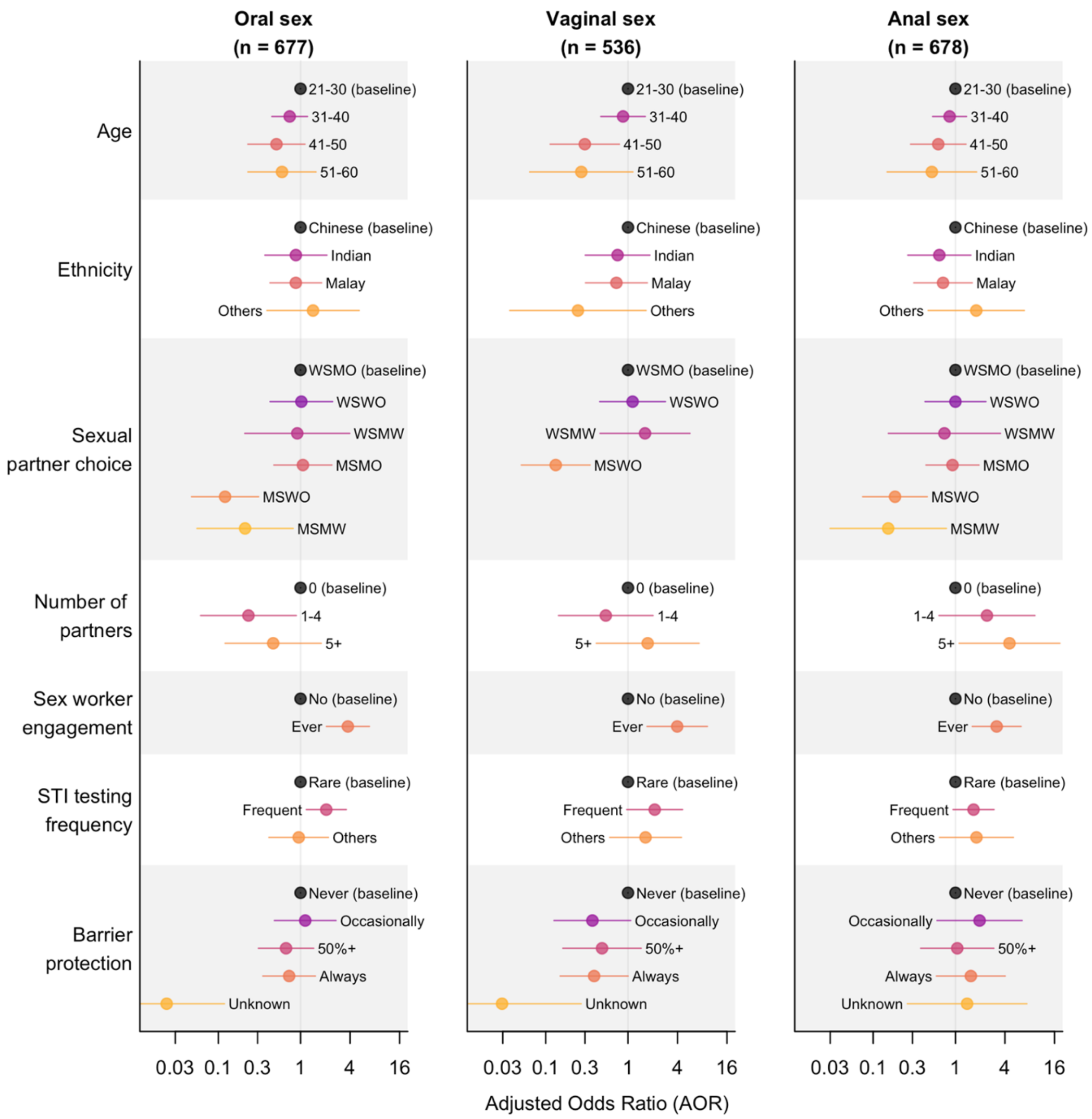

### Supplementary Figure 7

Adjusted odds ratio for socio-demographics, partnership-based sexual orientation defined by sex assigned at birth, number of sexual partners in the past year, engagement with sex workers, STI testing frequency, and barrier protection usage frequency, stratified by sexual activity type (columns). These include point estimates (dots) and corresponding 95% CIs (lines), quantifying the factors' impacts on STI positivity status. Extreme estimates due to insufficient samples (e.g., vaginal sex among MSMO) are not shown. Individuals who had never tested for STIs were excluded from the analysis. Rare STI testing is defined as testing once a year and frequent testing refers to testing once every three or six months. For barrier protection usage frequency, '50%+' indicates usage at least half of the time and 'always' means consistent use every time (Table S1). Please also note the x-axes are presented on a logarithmic scale.

### Fits for sexual activity patterns

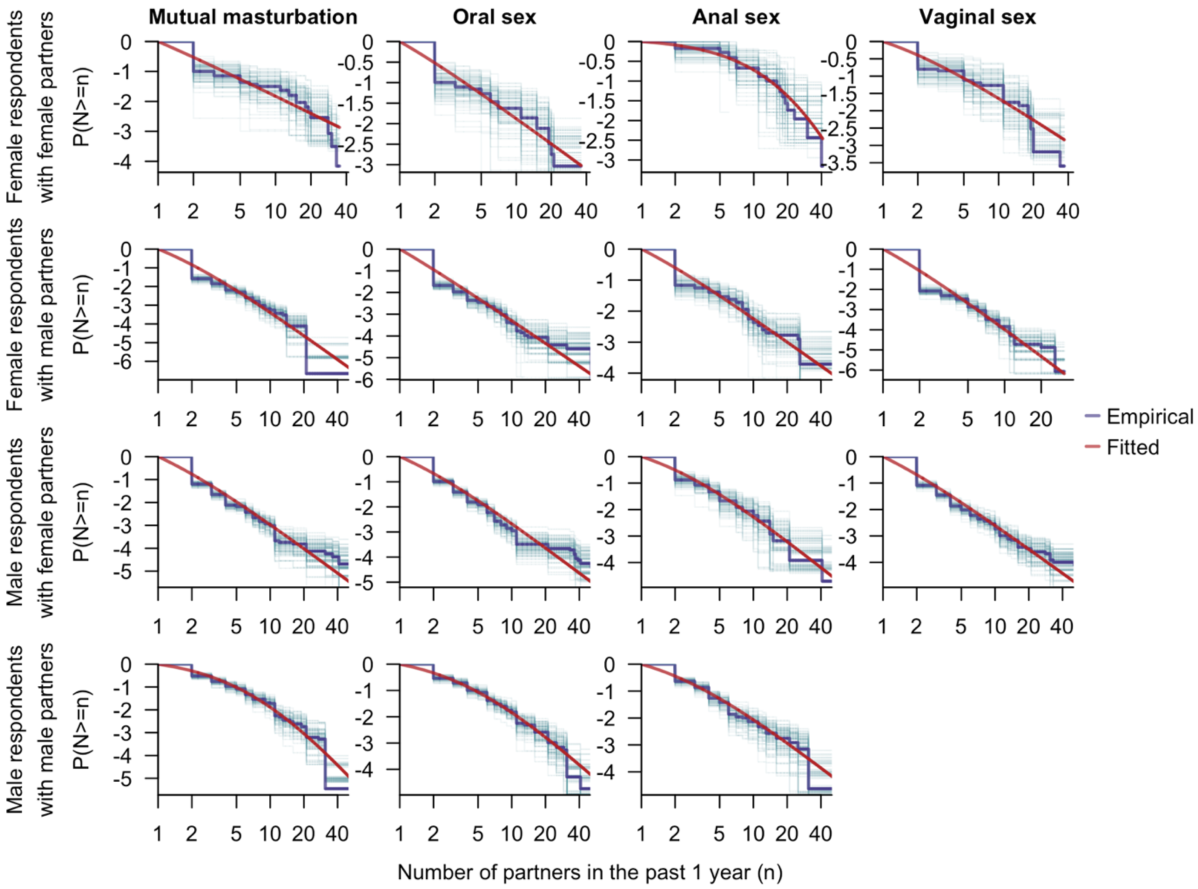

### Supplementary Figure 8

Distribution of the number of partners (N) in the past year among individuals with sexual partners, stratified by the sex assigned at birth of survey respondents and their partners (rows) and type of sexual activity (columns). The blue lines stand for the empirical distribution derived from bootstrap samples of the survey data, with the thick blue line in each subfigure indicating the pooled estimate. The red curves represent the fitted distributions. The distribution for the number of male partners among male respondents who engaged in vaginal sex with males was not fitted or presented due to the small sample size. Both the x- and y-axes are displayed on a logarithmic scale.

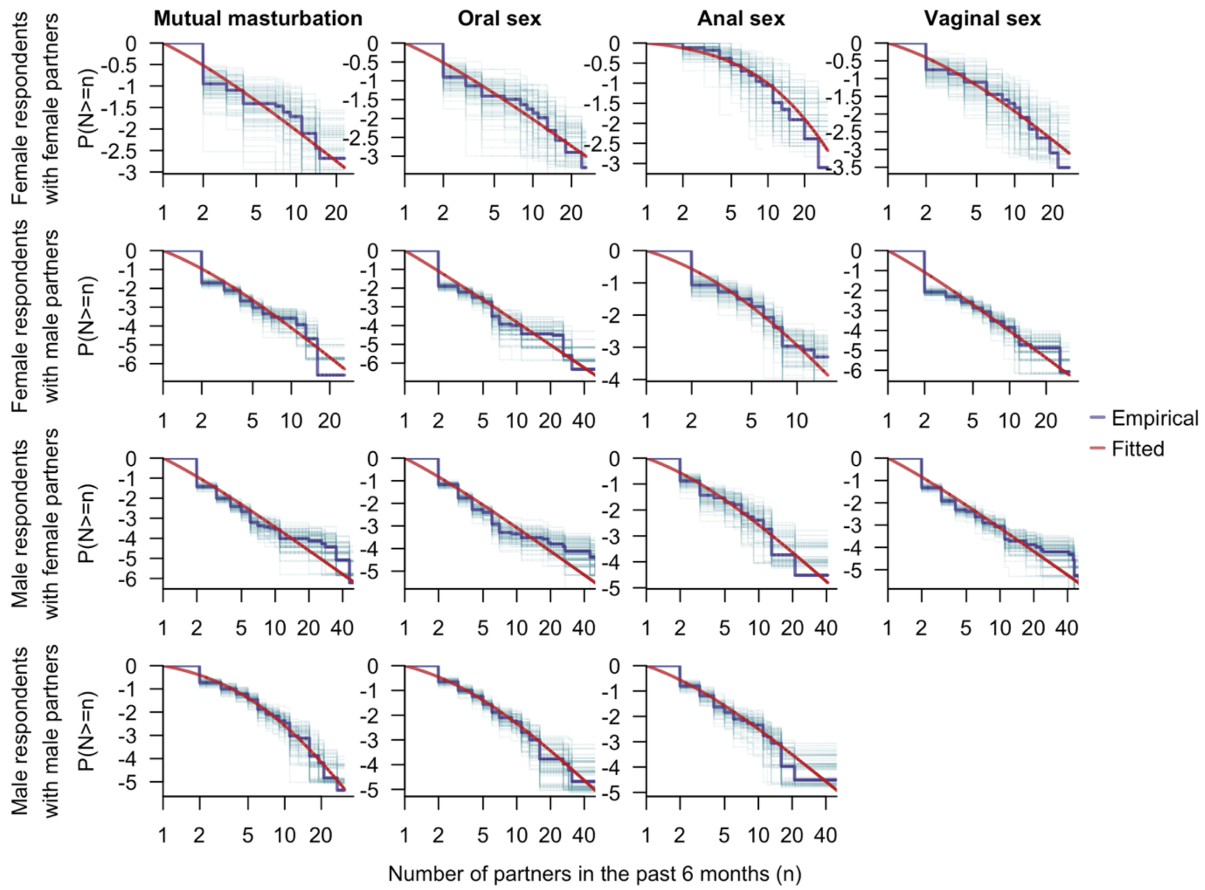

### Supplementary Figure 9

Distribution of the number of partners (N) in the past six months among individuals with sexual partners, stratified by the sex assigned at birth of survey respondents and their partners (rows) and type of sexual activity (columns). The blue lines stand for the empirical distribution derived from bootstrap samples of the survey data, with the thick blue line in each subfigure indicating the pooled estimate. The red curves represent the fitted distributions. The distribution for the number of male partners among male respondents who engaged in vaginal sex with males was not fitted or presented due to the small sample size. Both the x- and y-axes are displayed on a logarithmic scale.

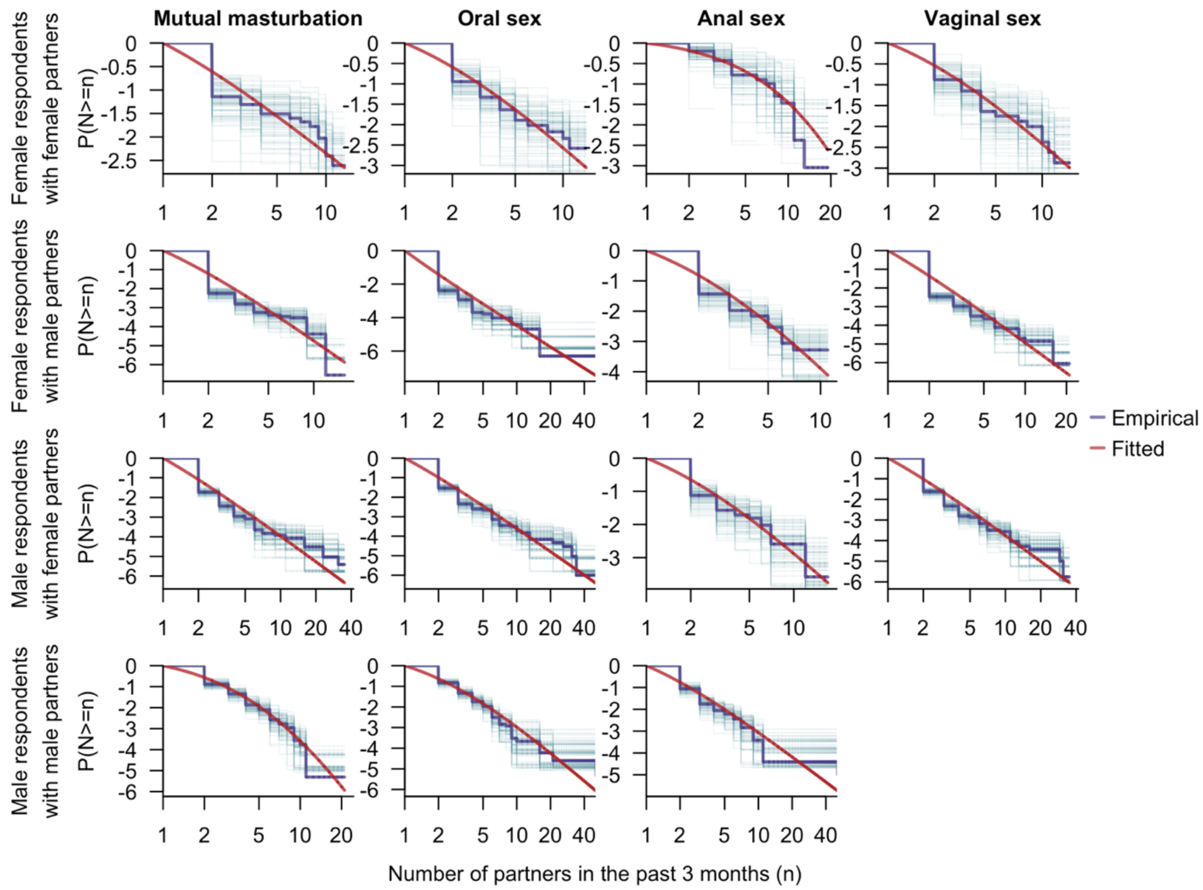

### Supplementary Figure 10

Distribution of the number of partners (N) in the past three months among individuals with sexual partners, stratified by the sex assigned at birth of survey respondents and their partners (rows) and type of sexual activity (columns). The blue lines stand for the empirical distribution derived from bootstrap samples of the survey data, with the thick blue line in each subfigure indicating the pooled estimate. The red curves represent the fitted distributions. The distribution for the number of male partners among male respondents who engaged in vaginal sex with males was not fitted or presented due to the small sample size. Both the x- and y-axes are displayed on a logarithmic scale.

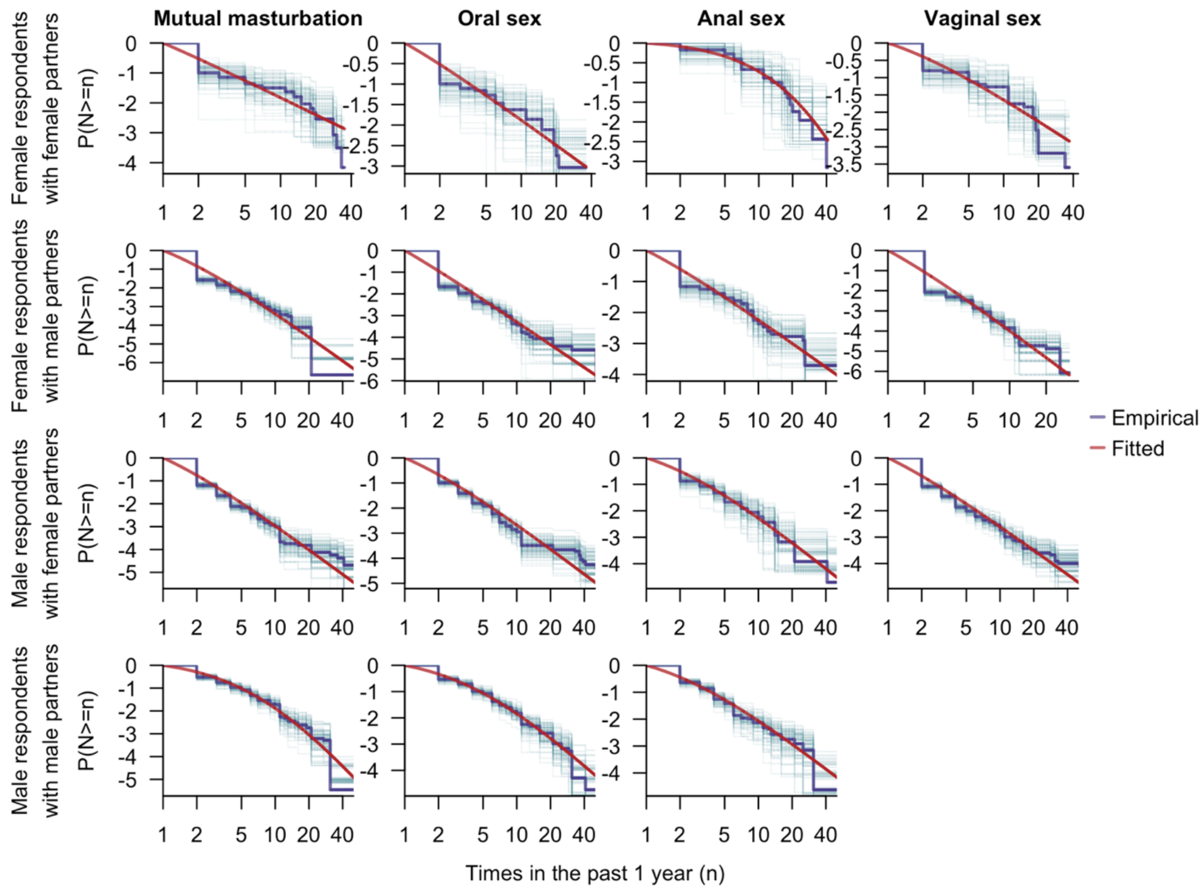

### Supplementary Figure 11

Distribution of the frequency of sexual activities (N) in the past year among individuals with sexual partners, stratified by the sex assigned at birth of survey respondents and their partners (rows) and type of sexual activity (columns). The blue lines stand for the empirical distribution derived from bootstrap samples of the survey data, with the thick blue line in each subfigure indicating the pooled estimate. The red curves represent the fitted distributions. The distribution for the number of male partners among male respondents who engaged in vaginal sex with males was not fitted or presented due to the small sample size. Both the x- and y-axes are displayed on a logarithmic scale.

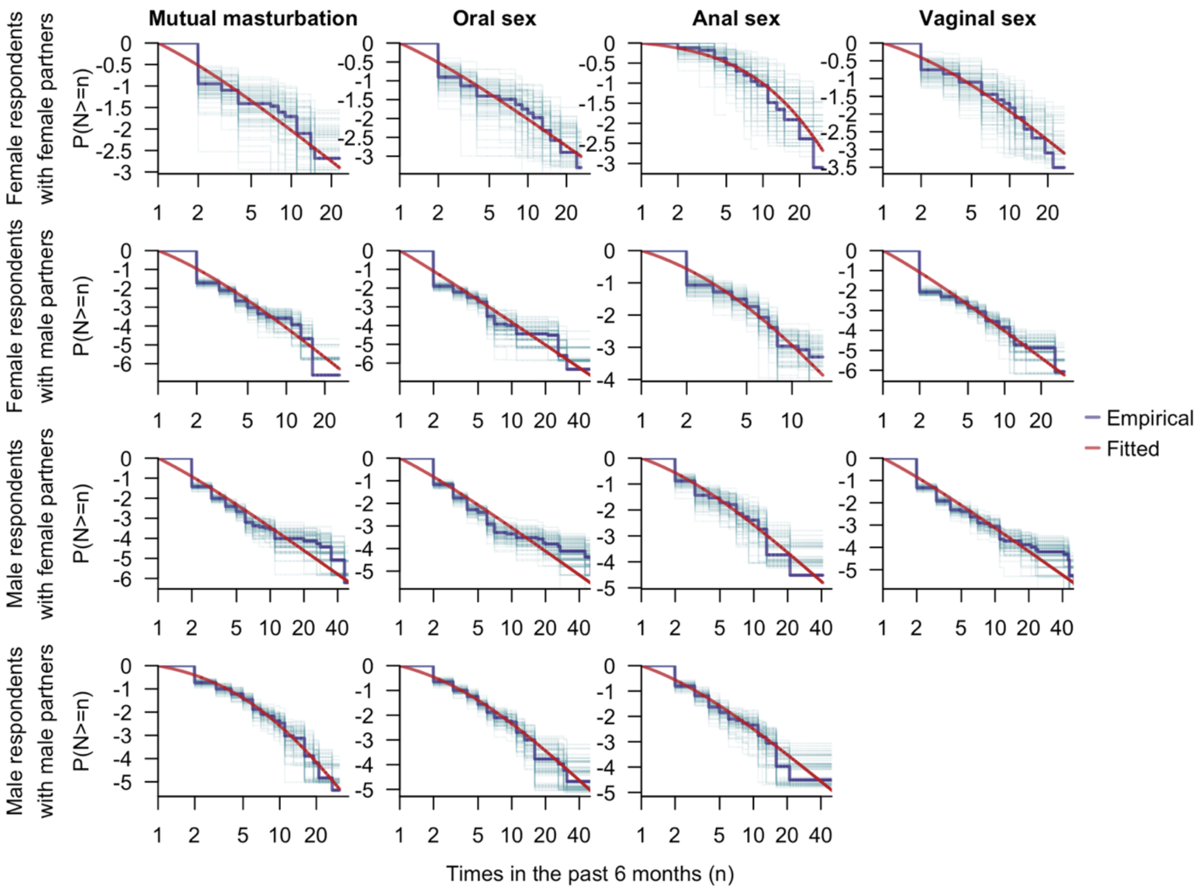

### Supplementary Figure 12

Distribution of the frequency of sexual activities (N) in the past six months among individuals with sexual partners, stratified by the sex assigned at birth of survey respondents and their partners (rows) and type of sexual activity (columns). The blue lines stand for the empirical distribution derived from bootstrap samples of the survey data, with the thick blue line in each subfigure indicating the pooled estimate. The red curves represent the fitted distributions. The distribution for the number of male partners among male respondents who engaged in vaginal sex with males was not fitted or presented due to the small sample size. Both the x- and y-axes are displayed on a logarithmic scale.

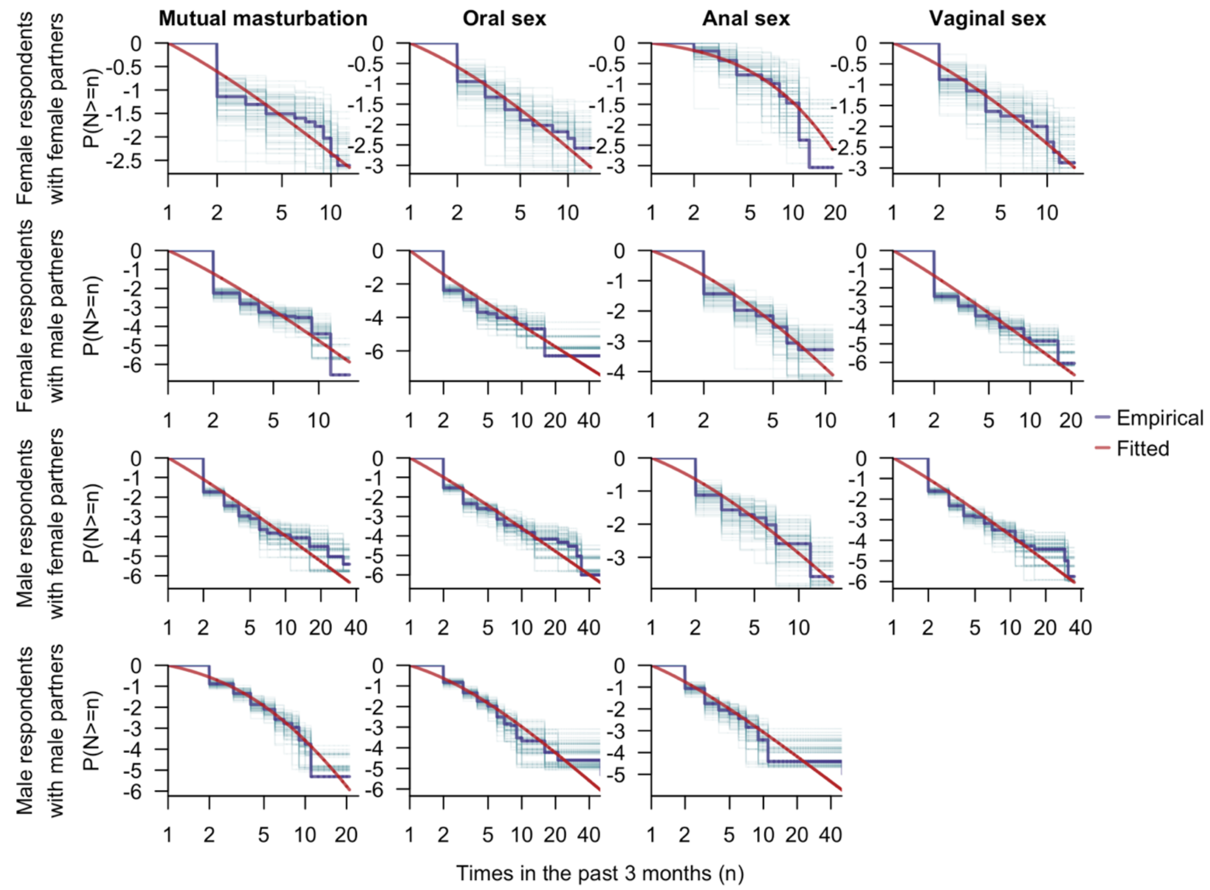

### Supplementary Figure 13

Distribution of the frequency of sexual activities (N) in the past three months among individuals with sexual partners, stratified by the sex assigned at birth of survey respondents and their partners (rows) and type of sexual activity (columns). The blue lines stand for the empirical distribution derived from bootstrap samples of the survey data, with the thick blue line in each subfigure indicating the pooled estimate. The red curves represent the fitted distributions. The distribution for the number of male partners among male respondents who engaged in vaginal sex with males was not fitted or presented due to the small sample size. Both the x- and y-axes are displayed on a logarithmic scale.

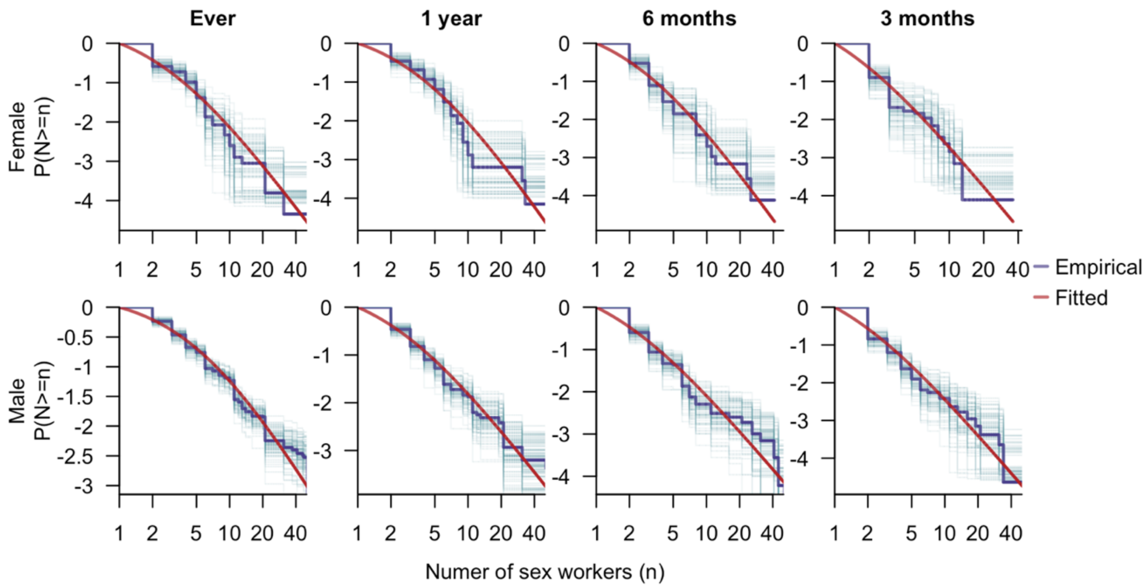

### Supplementary Figure 14

Distribution of number of sex workers (N) an individual has engaged with among those who reported engaging with sex workers, stratified by sex assigned at birth (rows) and recall period (columns). The blue lines stand for the empirical distribution derived from bootstrap samples of the survey data, with the thick blue line in each subfigure indicating the pooled estimate. The red curves represent the fitted distributions. The distribution for the number of male partners among male respondents who engaged in vaginal sex with males was not fitted or presented due to the small sample size. Both the x- and y-axes are displayed on a logarithmic scale.
